# Longitudinal peripheral blood transcriptional analysis of COVID-19 patients captures disease progression and reveals potential biomarkers

**DOI:** 10.1101/2020.05.05.20091355

**Authors:** Qihong Yan, Pingchao Li, Xianmiao Ye, Xiaohan Huang, Xiaoneng Mo, Qian Wang, Yudi Zhang, Kun Luo, Zhaoming Chen, Jia Luo, Xuefeng Niu, Ying Feng, Tianxing Ji, Bo Feng, Jinlin Wang, Feng Li, Fuchun Zhang, Fang Li, Jianhua Wang, Liqiang Feng, Zhilong Chen, Chunliang Lei, Linbing Qu, Ling Chen

## Abstract

COVID-19, caused by SARS-CoV-2, is an acute self-resolving disease in most of the patients, but some patients can develop a severe illness or even death. To characterize the host responses and identify potential biomarkers during disease progression, we performed a longitudinal transcriptome analysis for peripheral blood mononuclear cells (PBMCs) collected from 4 COVID-19 patients at 4 different time points from symptom onset to recovery. We found that PBMCs at different COVID-19 disease stages exhibited unique transcriptome characteristics. SARS-CoV-2 infection dysregulated innate immunity especially type I interferon response as well as the disturbed release of inflammatory cytokines and lipid mediators, and an aberrant increase of low-density neutrophils may cause tissue damage. Activation of cell death, exhaustion and migratory pathways may lead to the reduction of lymphocytes and dysfunction of adaptive immunity. COVID-19 induced hypoxia may exacerbate disorders in blood coagulation. Based on our analysis, we proposed a set of potential biomarkers for monitoring disease progression and predicting the risk of severity.

## INTRODUCTION

The recent global outbreak of COVID-19 is caused by a highly contagious new coronavirus named SARS-CoV-2 (*1-3*). WHO has declared the outbreak of COVID-19 a Public Health Emergency of International Concern (*4*). As of April 28, there have been more than 3 million confirmed cases and more than 210,000 deaths worldwide according to the reports of WHO. Most patients with COVID-19 showed mild or no severe symptoms. Fever, dry cough, dyspnea and ground hyaline pneumonia are the most common clinical manifestations. About 20% of patients develop severe disease or acute respiratory distress syndrome (ARDS). Clinical features include hypoxemia, lymphopenia, and thrombocytopenia (*5-8*).

In view of the rapid spread of COVID-19, lack of understanding of host response to SARS-CoV-2 has become a critical issue. Increased levels of serum proinflammatory cytokines, including IL-2, IL-7, IL-10, G-CSF, IP-10, MCP-1, MIP-1α, and TNF-α, are found in COVID-19 patients, which are higher in severe cases (*3*). Previous reports demonstrate that excessive proinflammatory cytokines (e.g., IFN-γ, IP-10, MCP-1, and IL-8) release is associated with pneumonia and lung damage in severe acute respiratory syndrome (SARS) and H5N1 patients (*9-11*). It has been reported that SARS-CoV-2 infection rapidly activates inflammatory T cells and inflammatory mononuclear macrophages through the GM-CSF and IL-6 pathways, leading to a cytokine storm and severe lung damage (*12*). SARS-CoV-2 infection causes a reduction in T cell numbers, which reduces the functional diversity of T cells in patients with COVID-19 (*13*). Functional exhaustion of NKG2A^+^ NK may be associated with disease progression in the early stages of COVID-19 (*14*). However, the innate and adaptive immune profiles and characteristics in COVID-19 patients during disease progression remain unclear.

To understand the host pathophysiological responses after SARS-CoV-2 infection, we performed a longitudinal analysis of transcriptomes for peripheral blood mononuclear cells (PBMCs) collected at 4 different time points between symptom onset and convalescent stage. In combination with laboratory tests and clinical observations, we identified potential biomarkers that may lead to better monitoring of the COVID-19 disease progression and for early prediction of prognosis.

## RESULTS

### PBMCs at different disease stages show distinct transcriptome signatures

We obtained a total of 16 blood samples 4 COVID-19 patients each with 4 different time points that ranged from early onset to convalescent stages (Fig. 1A). These patients were regular non-ICU cases and had mean hospitalization days of 21.5 ± 2.5 and mean SARS-CoV-2 positive days of 12 ± 1.8. The detailed information of these patients was described in Fig. S1A. Four blood samples from a healthy donor before and after vaccination with a QIV inactivated seasonal influenza virus vaccine were used as healthy controls. RNA sequencing (RNA-seq) using Illumina HiSeq3000 was performed at the same time for 20 samples of 2 to 4 million PBMCs. A total of 2.2 billion reads or an average of 102 million reads per sample was obtained after quality control processing (Fig. S1B). The transcripts of SARS-CoV-2 virus receptors ACE2 and TMPRSS2 were undetectable or extremely low in PBMCs (Fig. S1C). No fragments of SARS-CoV-2 viral genome could be found in all samples (Fig. S1B), suggesting that SARS-CoV-2 does not significantly infect human PBMCs, at least in non-severe cases.

**Fig. 1.**
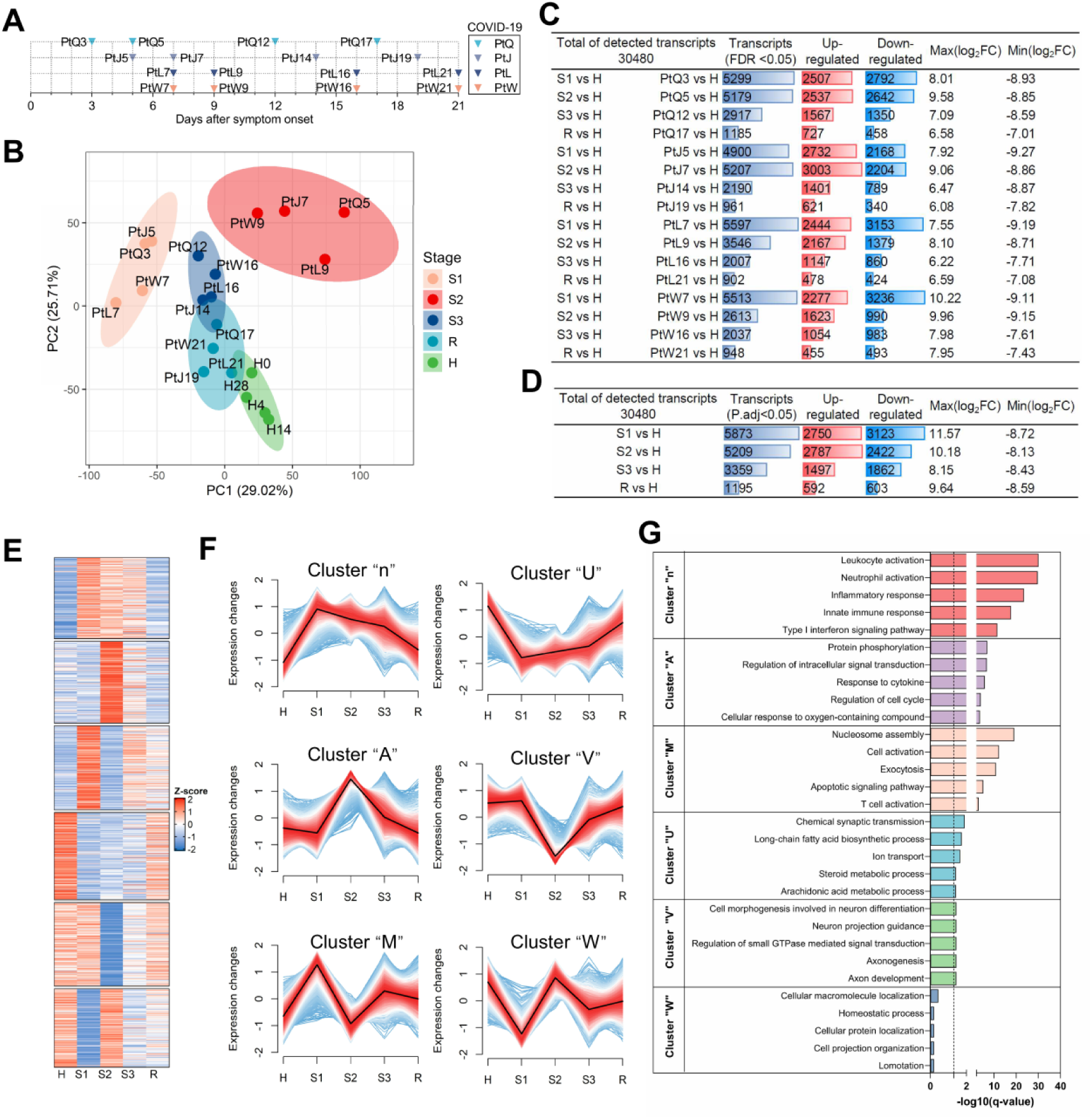
Global transcriptional analysis across COVID-19 patients and health donor samples. (**A**) Time points of blood sample collection from 4 COVID-19 patients. (**B**) Principal component analysis in COVID-19 patients and health donor samples, depicting the variation in the global gene expression profiles across different stages (S1, S2, S3, and R) and healthy control (H). Principal components 1 (PC1) and 2 (PC2), which represent the greatest variation in gene expression, are shown. (**C**) Paired comparison between S1 and R, S2 and R, S3 and R for each COVID-19 patient. The numbers of up-regulated and down-regulated gene are listed. (**D**) Grouped comparison between S1 and R, S2 and R, S3 and R. Samples from different patients in the same stage were combined and compared with samples from the convalescent stage. The numbers of up-regulated and down-regulated gene are listed. (**E**) All DEGs were grouped into six clusters according to their expression pattern. Heatmap showing the relative expression of individual transcript at different disease stages. (**F**) The relative expression changes of each cluster are shown. Each line in the plots presents a unique DEG and the black line indicates the median. (**G**) Gene ontology (Go) analysis for each cluster. Top 5 Go terms enriched in each cluster are shown. The dotted line indicates the threshold value (q-value = 0.05) of significantly enriched.

Principal component analysis (PCA) of gene expression grouped the patient samples into 4 clusters. Interestingly, these 4 clusters coincided with disease progression in clinical observation (Fig. 1B, Fig. S1A). We named these four clusters as: 1) Stage 1 (S1), representing the early onset; 2) Stage 2 (S2), representing the clinically most severe stage; 3) Stage 3 (S3), representing improving stage; and 4) Stage R, representing the recovering or convalescent stage. Notably, all three patients in S2 (PtQ5, PtJ7, PtL9, PtW9) were at the most severe disease state, demonstrated by the highest C-reactive protein in plasma, the lowest lymphocyte counts, and the worst chest radiography (Fig. S1A). Four samples from the healthy donor formed a distinct cluster, which was named as cluster H. Of note, cluster R is adjacent to cluster H, suggesting the transcriptomes in the recovery stage is approaching the healthy state. This result demonstrates that the gene expression profiles have distinct patterns along with disease progression. These patterns were reproducible in different COVID-9 patients, at least for non-ICU patients.

We performed a digital cytometry CIBRSORTx (*15*) to delineate the transcriptome into abundances of cell subsets (Fig. S1D) in the PBMCs at different stages of disease progression. This analysis showed a dramatic increase of monocytes and pathological low-density neutrophils in peripheral blood during S1 and S2 (Fig. S1D). In contrast, there was a reduction of T and NK cells in S1 and especially S2. The perturbation in the proportion of cell types in PBMCs, especially lymphocytopenia is one of the leading clinical manifestations of COVID-19. Our analysis was verified by clinical tests of T lymphocyte and neutrophil counts in these patients (Fig. S1A).

### Time series-based global gene expression pattern analysis of DEGs reveals three main clusters during the disease progression

To identify the differentially expressed genes (DEGs) at different disease stages, we first performed paired comparison between S1 and R, S2 and R, S3 and R for each person (Fig. 1C). We also combined all samples from different patients at the same disease stage for group comparison (Fig. 1D). Compared to the convalescent state, the number of DEGs was 2661, 4811, and 834 in S1, S2, and S3 respectively, representing 8.7%, 15.8%, and 2.7% genes in the transcriptomes (Fig. 1D). In contrast, there were few changes in the number of DEGs before and after vaccination in a healthy person. We found that there were many identical DEGs among different COVID-19 patients, especially when these samples belonged to the same disease stage. The 4 patients shared 155, 341 and 23 DEGs at S1, S2 and S3 respectively (Fig. S1E-F). Therefore, SARS-CoV-2 infection resulted in common changes in gene expression profile in different regular COVID-19 patients. Since the samples in R may not fully represent the healthy state, we also performed grouped comparison between S1 and H, S2 and H, S3 and H, R and H (Fig. S1G). Indeed, there were more DEGs when compared with gene expression in a healthy state. An average of 19.3%, 17.1%, 11.0%, and 3.9% genes in the transcriptome have undergone significant expression changes in S1, S2, S3, and R, respectively.

To identify genes closely related to disease progression, we performed a time series analysis for all DEGs. The healthy samples were assumed to be a stage representing pre-infection to capture the time-dependent changes. All DEGs could be divided into 6 clusters based on their expression patterns (Fig. 1E). The pattern of gene expression in each cluster somewhat appeared to resemble an alphabet, so we named each cluster as “n”, “A”, “M”, “U”, “V”, and “W”. Each cluster contains 1659, 1668, 1717, 1780, 1703, and 1599 genes respectively (Fig. 1F). To investigate the biological functions associated with each cluster, we performed gene ontology analysis. The top 5 biology process (BP) terms of each cluster were listed (Fig. 1G). Cluster “n”, “A”, and “M” contained 1043, 609, and 339 BP terms respectively (q-value <0.05). Genes in cluster “U”, “V” and “W” showed few discernible biological themes; each contained only 23, 11, and 0 BP terms (Fig. 1G, Fig. S1H). To verify the contribution of the genes in cluster “n”, “A” and “M” to disease progression, we took the top 2000 genes with the largest contribution to PC1 and PC2 (termed gene set PC1 and PC2 hereafter) and compared with genes in the cluster “n”, “A” and “M”, respectively (Fig. 1B, Fig. S1I). The result revealed that genes in cluster “n” highly overlapped with gene set PC2, which reflects the difference between illness and healthy state. Genes in cluster “M” highly overlapped with gene set PC1, which reflects disease severity. Genes in cluster “A” partially overlapped with both PC1 and PC2. Therefore, we mostly focused on these three clusters (n, A and M) that were likely associated with disease progression in the subsequent analysis.

### Inflammatory cytokines and lipid mediators are disturbed in COVID-19 patients

We noticed that a set of DEGs in cluster “n” were enriched in the biological processes: immune effector process, response to cytokine, inflammatory response, cytokine production, and cell chemotaxis (Fig. 2A-B). Their expression increased in S1 and S2, declined in S3 and R (Fig. 2C, Fig. S2A). These “n” type genes include: IL1B, IL6, IL10, IL12A, IL18, IL19, IL27, and chemokine CXCL10 (IP-10), are known to promote the inflammatory response. Persistent high-level expression of chemokines CXCL1, CXCL2, CXCL3, and CXCL8 promote neutrophil infiltration into the lung and/or other tissues, while CCL2 promotes monocyte migration to the site of infection (Fig. 2C, Fig. S2A). Interestingly, we found another set of genes in cluster “A”, including cytokines such as IL2, IL7, IL15, and IL21, their expression sharply elevated in S2 and returned to near-normal levels, as clinical symptoms resolved in S3 and R (Fig. 2C, Fig. S2A). These cytokines are associated with the proliferation and activation of T, B, and NK cells. To verify our analysis, we used Luminex to confirm the protein levels of some cytokines in the plasma of COVID-19 patients in this study. IP-10 and IL-6 were indeed significantly elevated in S1 and S2, but returned to near normal range in S3 and R as the disease starting to resolve (Fig. 2D). These findings further indicated that dysregulated cytokines release strongly associated with the progression of the disease.

**Fig. 2.**
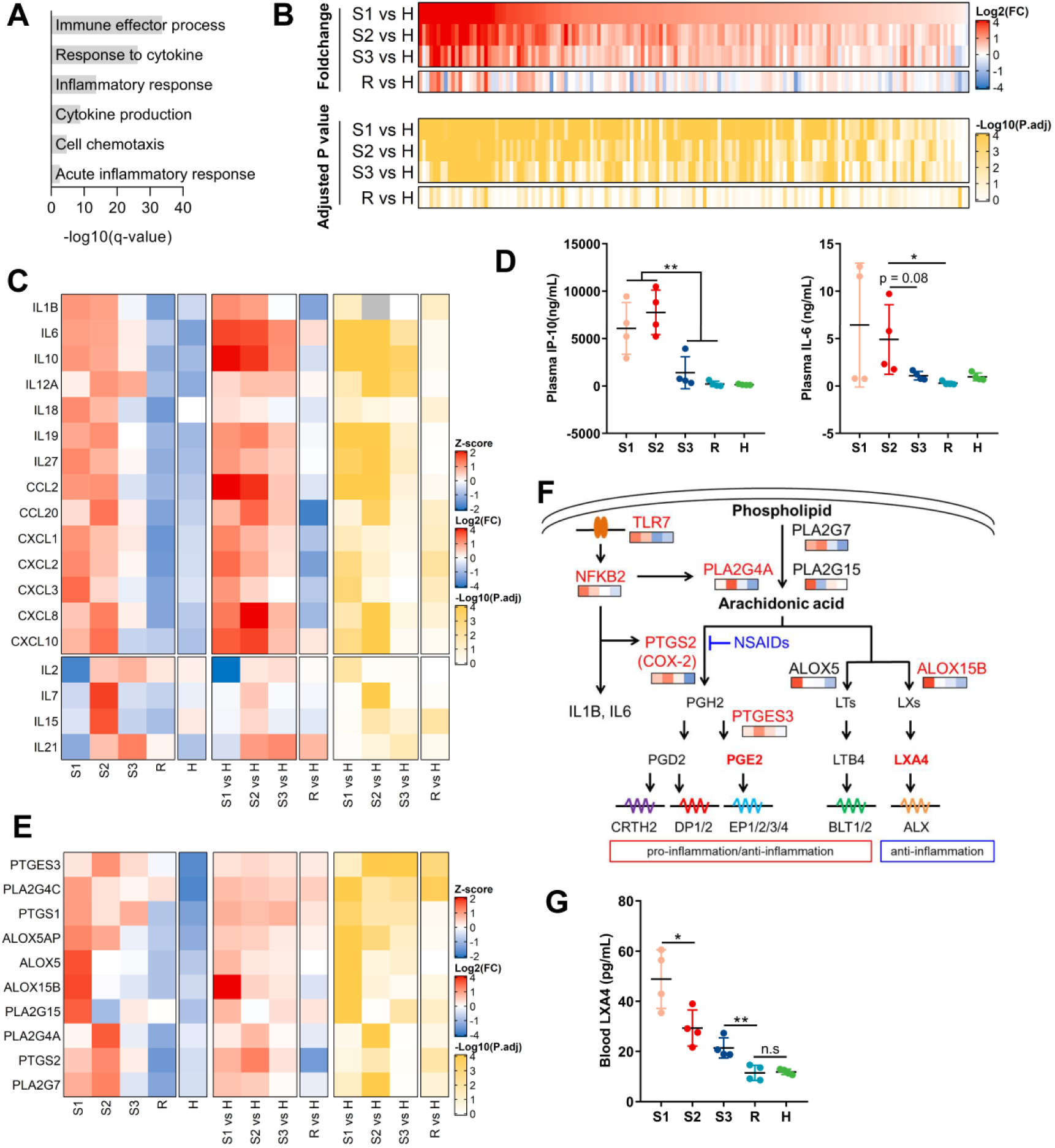
Dysregulated inflammatory cytokines and lipid mediators in COVID-19 patients. (**A**) Gene ontology (Go) analysis of transcripts included in cluster “n” showing enrichment for cytokine-related biological processes. Horizontal axis denotes statistical significance as measured by minus logarithm of q-values. Vertical axis ranked the Go terms by q-values (gray bars). (**B**) Heatmaps showing foldchange (top panel) and corresponding adjusted p values (bottom panel) for cytokine transcripts. (**C**) Heatmaps showing relative expression level (left panel), foldchange (middle panel) and adjusted p values (right panel) for a set of selected inflammatory cytokines and chemokines. (**D**) Plasma levels of IP-10 and IL-6 among healthy controls and COVID-19 patients in four stages. **, p<0.01; *, p<0.05 by paired t test. (**E**) Heatmaps showing relative expression level (left panel), foldchange (middle panel) and adjusted p values (right panel) for a set of genes involved in prostanoids synthesis. (F) Schematic representation prostanoids synthesis pathways. The expression pattern of enzymes involved in prostanoids synthesis is showed under gene names. PLA2s catalyze membrane phospholipid to release AA, which is converted to PGH2 by COX-1 (PTGS1) and COX-2 (PTGS2). PGH2 is then converted by PTGDS and PTGES to PDG2 and PGE2. AA is also converted to leukotrienes and lipoxins by 5-LO (ALOX5) and 15-LO (ALOX5). (G) Plasma levels of LXA4 among healthy controls and COVID-19 patients in four stages. **, p<0.01; *, p<0.05; n.s, p>0.05 by paired t test.

We found that lipid mediators also play an important role in the inflammatory response of COVID-19 patients. The expression pattern of enzymes involved in lipid mediator synthesis also belonged to cluster “n”, including cyclooxygenases and lipoxygenases which convert arachidonic acid (AA) to lipid mediator prostanoids. The increased expression of TLR7 and NFKB2 promoted the expression of PLA2G4A and PTGS2 (COX-2) in S1 (Fig. 2E). The key enzymes of lipid mediator synthesis, including PLA2G7, PLA2G15 and PTGES3 (PGE2 synthase), also increased in S1 and S2 (Fig. 2E-F, Fig. S2B). Elevation of these enzymes can promote the production of inflammatory prostanoids and inflammatory response. Interestingly, lipoxygenase ALOX15B, which produces anti-inflammatory mediator LXA4, had a surge in S1 as an initial response, but declined in S2 and resumed to near healthy state in R as the disease resolved (Fig. 2E-F, Fig. S2B). We measured the plasma concentration of LXA4 in these patients and found that LXA4 also increased significantly in S1 and then declined in S2 and thereafter (Fig. 2G). Therefore, lipid mediators are likely to play an important role in inflammation-associated disease severity.

### Elevated pathological low-density neutrophils may contribute to tissue damage

We found that neutrophil-associated genes were highly enriched in cluster “n” (Fig. 3A-B). During viral infections or autoimmune diseases, neutrophils may abnormally differentiate to pathological low-density neutrophils (LDNs) that remain in PBMCs after Ficoll-gradient centrifugation (*16, 17*). Our digital cytometry showed an increase of low-density neutrophils during S1 and S2 (Fig. S1D, Fig. 3D). Besides, complete blood count verified that there was an elevated neutrophils level in S2 in these patients (Fig. 3E). A general characteristic of LDNs is the simultaneous expression of neutrophils activation markers FCGR3B (CD16b), CEACAM8 (CD66b), and ITGAM (CD11b), and neutrophils differentiation markers typically expressed in immature granulocytes (LRG1). The expression of these genes significantly increased in S1 and S2, and gradually returned to near normal in S3 and R (Fig. 3C, Fig. S3A). These observations indicated that COVID-19 could promote the development of LDNs, which is associated with disease progression.

**Fig. 3.**
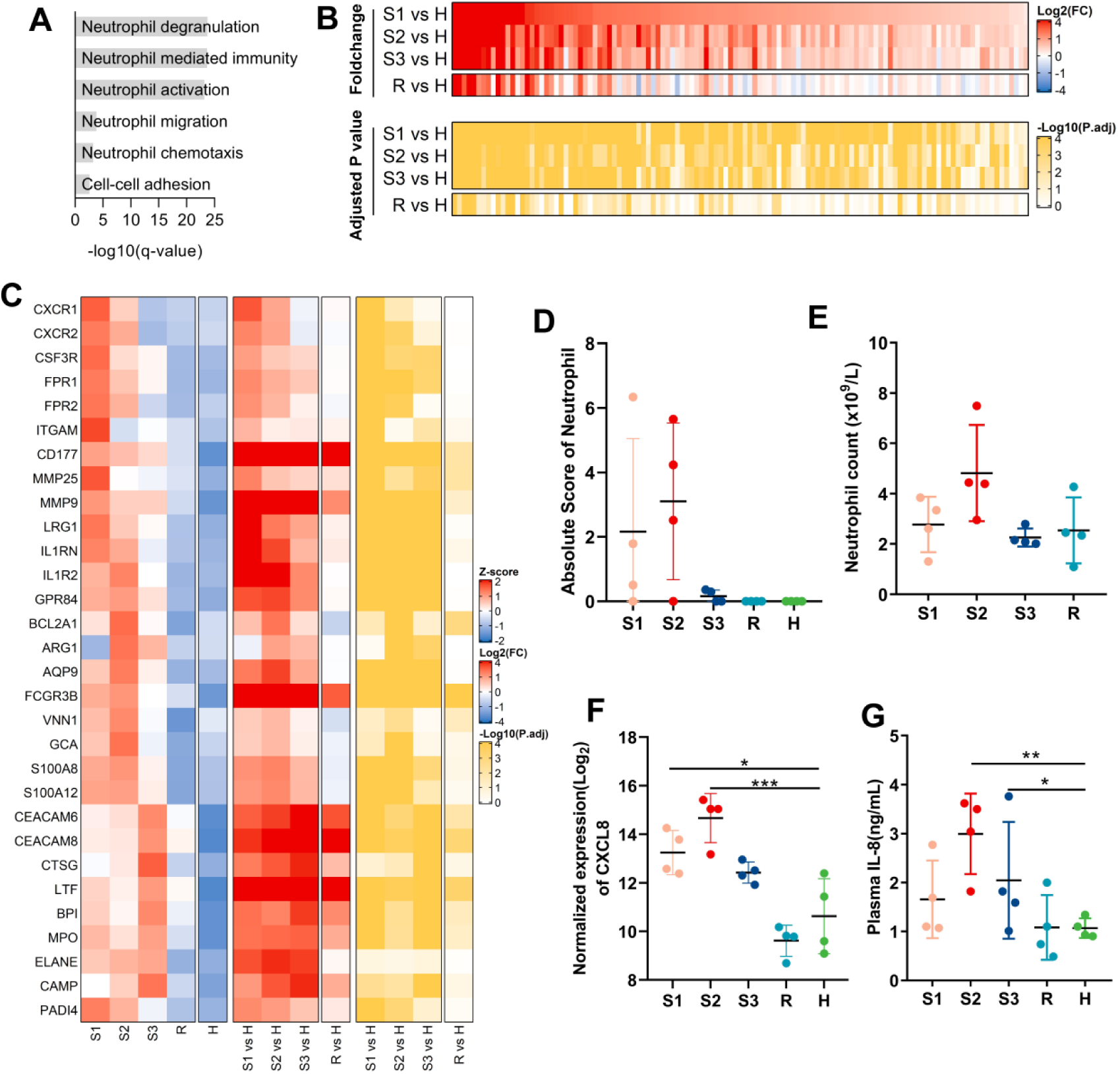
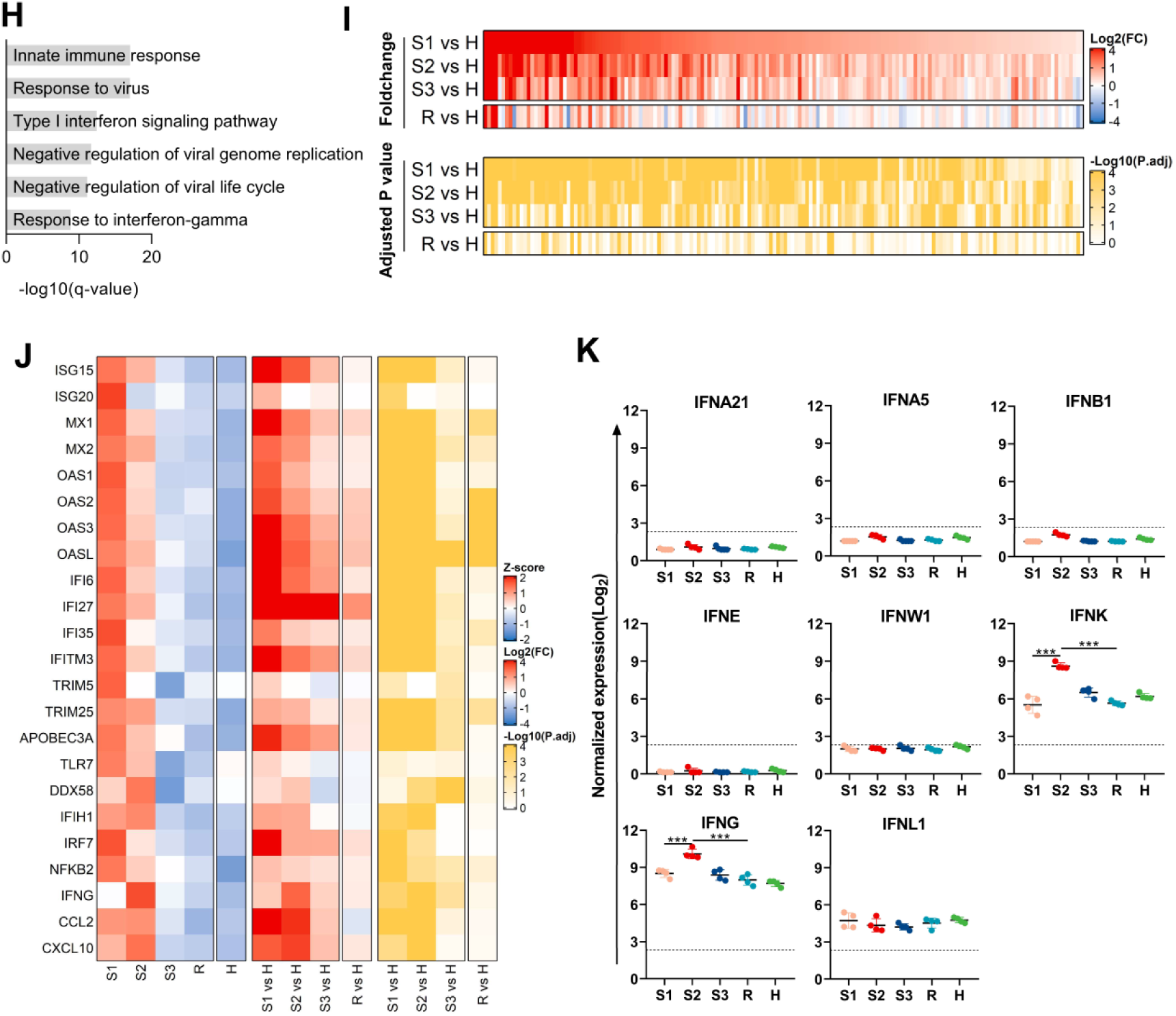
Abnormal neutrophils and imbalanced interferon responses in COVID-19 patients. (**A**) Go analysis of transcripts included in cluster “n” showing enrichment for neutrophil-related biological processes. Horizontal axis denotes statistical significance as measured by minus logarithm of q-values. Vertical axis ranked the Go terms by q-values (gray bars). (**B**) Heatmaps showing foldchange (top panel) and corresponding adjusted p values (bottom panel) for neutrophils-related transcripts. (**C**) Heatmaps showing relative expression level (left panel), foldchange (middle panel) and adjusted p values (right panel) for a set of selected neutrophils-related transcripts. (**D**) Absolute neutrophil abundance derived using the CIBERSORTx algorithm. (**E**) Absolute neutrophil counts in peripheral blood determined by complete blood count. (**F**) Normalized log_2_ expression of CXCL8 (encoding IL-8) from individual COVID-19 patients or healthy controls. ***, p<0.001; **, p<0.01; *, p<0.05 by paired t test. (**G**) Plasma IL-8 concentration of healthy controls and COVID-19 patients in four stages. **, p<0.01; *, p<0.05 by paired t test. (**H**) Go analysis of transcripts included in cluster “n” showing enrichment for innate immune-related biological processes. Horizontal axis denotes statistical significance as measured by minus logarithm of q-values. Vertical axis ranked the Go terms by q-values (gray bars). (**I**) Heatmaps showing foldchange (top panel) and corresponding adjusted p values (bottom) for innate immune-related transcripts. (**J**) Heatmaps showing relative expression level (left panel), fold change (middle panel) and adjusted p values (right panel) for a set of selected innate immune-related transcripts. (**K**) Normalized log_2_ expression of IFNA5, IFNA21, IFNB1, IFNE, IFNW1, IFNG, and IFNL1 from individual COVID-19 patients or healthy controls. The dotted line represents the detection line. ***, p<0.001; **, p<0.01; *, p<0.05 by paired t test.

LDNs exhibit enhanced capacity to release neutrophil extracellular traps (NETs), which are composed of cellular DNA, core histones, and azurophilic granule proteins. Excessive NETs release or impaired NETs clearance causes tissue damage. The key enzyme (PADI4) and azurophilic granule proteins (MPO, CTSG, MMP9 and ELANE) of NETs formation in COVID-19 increased in S1, S2, and S3. As clinical symptoms alleviated in R, these genes declined to near-normal levels (Fig. 3C, Fig. S3A). The “n” cluster also contains genes related to neutrophil chemotaxis and migration. CXCR1, CXCR2, FPR1, FPR2, CD177 and AQP9 are known to regulate neutrophil chemotaxis, which also increased in S1 and S2 (Fig. 3C, Fig. S3A). Furthermore, the transcripts of inflammatory-related receptors (GPR84, IL1R2) also elevated in S1, S2, and S3 (Fig. 3C, Fig. S3A). CXCL8 (IL-8) is secreted primarily by neutrophils. Its elevation in S1 and S2 (Fig. 3F) may acts as a chemotactic factor by further recruiting the neutrophils to the site of infection. We measured the plasma concentration of IL-8 in these patients and found that plasma IL-8 started to increase in S1, peaked in S2, decreased in S3, and returned to normal level in R (Fig. 3G). Therefore, the increase of LDNs may contribute to pneumonia and tissue damage.

### The interferon responses are imbalanced in COVID-19 patients

We found that the genes involved in innate immunity process were enriched in cluster “n” (Fig. 3H-I). The single-stranded RNA sensors TLR7, DDX58 (RIG-I), IFIH1 (MDA5) and their downstream transcription factors IRF7 and NFKB2 increased in S1-S2, sharply decreased in S3, and gradually reached to near normal level in R (Fig. 3J, Fig. S3B). Interestingly, several subtypes of type I IFNs, such as IFNA (IFN-α), IFNB (IFN-β), IFNE (IFN-ε) and IFNW (IFN-ω) was almost undetectable from S1 to R (Fig. 3K). In contrast, another subtype of type I IFNs, IFNK (IFN-κ), and type II IFNs, IFNG (IFN-γ), started to increase in S1, peaked in S2, decreased in S3 and reached the normal level in R (Fig. 3K). While type III IFNs, IFNL (IFN-λ) was unchanged from S1 to R (Fig. 3K). These results indicate that the expression of interferons is dysregulated in COVID-19 patients.

Nevertheless, there was a significant increase in type I interferon-stimulated genes (ISGs) in S1, including ISG15, ISG20, TRIM5, TRIM25, and APOBEC3A, IFN-induced GTP-binding protein (MX1 and MX2), 2’-5’-oligoadenylate synthase (OAS1, OAS2, OAS3, and OASL), and interferon-inducible protein (IFI6, IFI27, IFI35, and IFITM3). These ISGs decreased in S2, and gradually reduced to normal level in S3 and R (Fig. 3J, Fig. S3B). These results suggested that the antiviral response was significantly enhanced in early infection. Interestingly, the IFN-γ inducible genes CCL2, CXCL10 showed similar expression patterns as IFNG, which increased in S1-S2, decreased in S3-R (Fig. 2C, 3J-K). These results demonstrated that the significant elevation of type II interferon and type II interferon inducible cytokines might be a hallmark that the disease is in the most severe stage.

### COVID-19 patients exhibit dysregulated adaptive immune responses

Adaptive immunity plays a critical role in the clearance of virus. We found that most genes involved in T cell and B cell responses were enriched in cluster “M” (Fig. 4A, B). T cell markers and proximal signaling molecules, CD3E (CD3ε), CD3G, CD4, CD8A, CD8B and LCK, ZAP70, CD247 (CD3ξ), LAT, GRB2, VAV1, CD6, increased in S1, declined to the lowest level in S2, then elevated in S3 and slightly decreased again in R (Fig. 4C, Fig. S4A). Expression of TCR was at the lowest level in S2, and rebounded in S3-R (Fig. 4D). Interestingly, the decrease of TCR diversity occurred in S1, remain low in S2, increased in S3 and R, but was still much lower than the healthy state (Fig. 4E), suggesting T-cells started to return to circulation in S3 and R. Of note, GZMB (granzyme B) increased in S1, but decreased in S2, then increased again in S3-R, indicating CD8^+^ T cells may undergo hyperactivation in response to viral infection during early-onset (Fig. 4C, Fig. S4A). The molecules highly expressed by B cells, including CD19, IL4R, PAX5, MZB1, XBP1, PRDM1, IRF4,TNFRSF13B (TACI), TNFRSF13C (BAFFR), CD79A, CD79B and the markers of antibody-secreting cells (ASCs), CD38 and CD27 elevated in S1, reduced in S2, increased in S3, and then decreased in R (Fig. 4F, Fig. S4B). BCR expression increased in S1, decreased in S2, increased in S3, then decreased again in R (Fig. 4G). BCR diversity was at the lowest level in S2, elevated in S3-R, and peaked in R (Fig. 4H). Therefore, B cells underwent activation, clonal selection and expansion in responding to exposure to viral antigens. Of note, we found that two most abundant antibody heavy chain and light chain transcripts, IGHV3-9 and IGLL5, had the same expression pattern (Fig. 4F, Fig. S4B), suggesting B cells differentiated into ASCs to secrete antibodies during the very early infection. Our analysis was supported by the peripheral blood counts of these patients, showing the lowest lymphocytes in S2 (Fig. 4I). We performed a quantitative RT-PCR which confirmed that CD4 and CD8 transcripts were at the lowest in S2 (Fig. 4J). Collectively, these results indicated that there were a dysregulated T cell and B cell responses during early onset but can recover as the disease resolved.

**Fig. 4.**
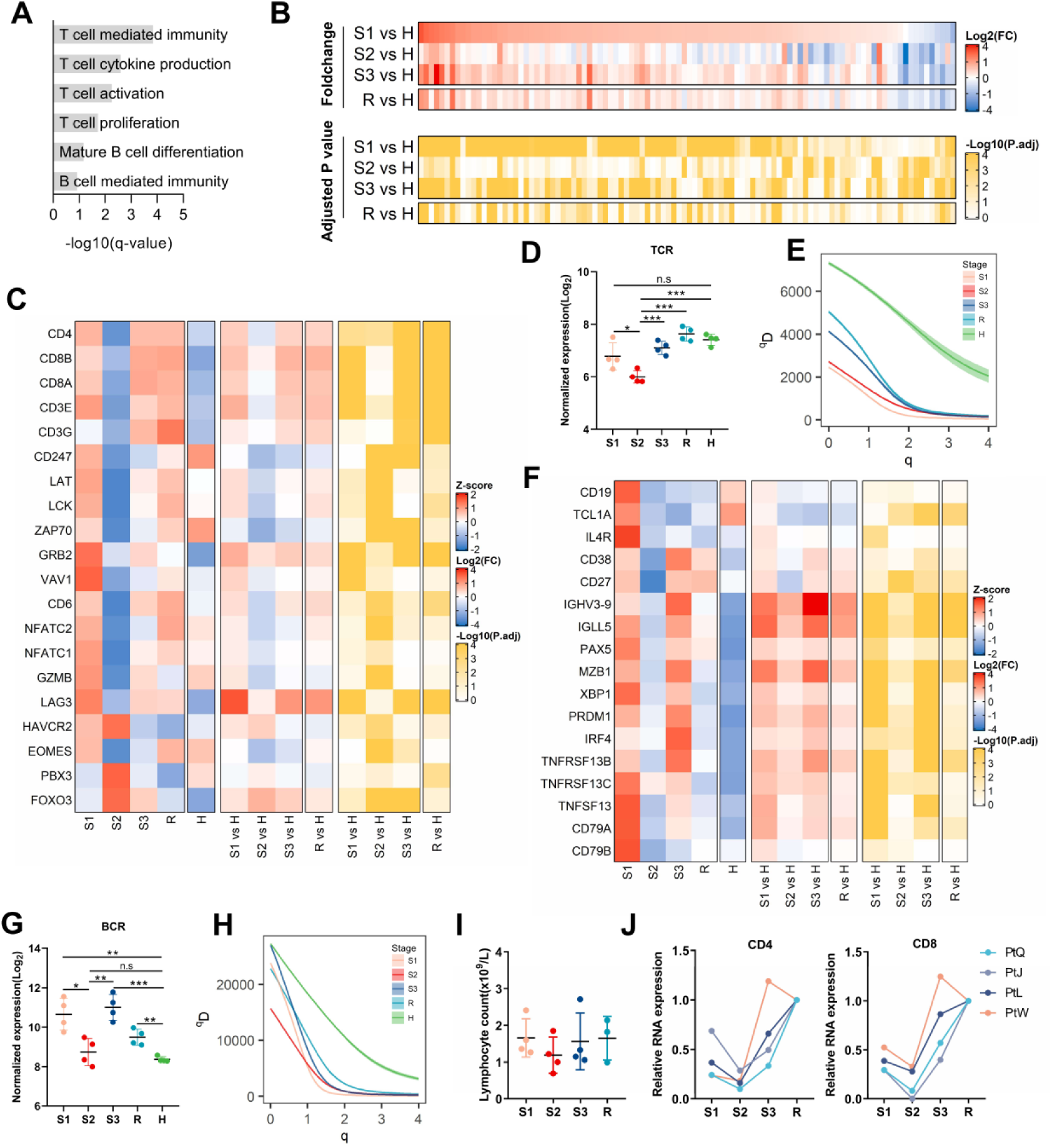
Dysregulated adaptive immune responses induced by SARS-CoV-2 infection. (**A**) Go analysis of transcripts included in cluster “M” showing enrichment for T and B cell response. Horizontal axis denotes statistical significance as measured by minus logarithm of q-values. Vertical axis ranked the Go terms by q-values (gray bars). (**B**) Heatmaps showing foldchange (top panel) and corresponding adjusted p values (bottom panel) for adaptive immune-related transcripts. (**C**) Heatmaps showing relative expression level (left panel), foldchange (middle panel) and adjusted p values (right panel) for a subset of T cell associated genes. (**D**) Normalized log2 expression of TCR for each sample from individual COVID-19 patients or healthy controls. (**E**) TCR diversity curve. Comparison of the Hill diversity index (qD, y-axis) over varying diversity orders *q* (x-axis, see Method), between each stage. (**F**) Heatmaps showing relative expression level (left), foldchange (middle) and adjusted p values (right) for a subset of B cell associated genes. (**G**) Normalized log_2_ expression of BCR for each sample from individual COVID-19 patients or healthy controls. (**H**) BCR diversity curve. Comparison of the Hill diversity index (*qD*, y-axis) over varying diversity orders *q* (x-axis, see Method), between each stage. (**I**) Lymphocyte counts for each sample from individual COVID-19 patients. (J) Relative expression of CD4 and CD8 by quantitative RT-PCR.

### Activation of cell death, exhaustion, and migration contribute to a reduction of lymphocytes and NK cells

We found that genes involved in cell death pathway were enriched in several clusters (Fig. 5A, B). Notably, many pro-apoptotic molecules were enriched in cluster “M”, which was similar to T and B cell-related genes. TP53 (p53), BAX, BAK1, BAD, BIK increased in S1, decreased in S2, elevated in S3, and then decreased to normal level in R. The extrinsic apoptosis, necrotosis, and pyroptosis related genes TNFRSF12A (TWEAK), TNFSF10 (TRAIL), FADD, CASP3, CASP7, CASP9, CYCS (cytochrome c), RIPK1, MLKL, and GSDME, also elevated in S1 or S2 (Fig. 5C, Fig. S5A). In contrast, the anti-apoptotic gene BCL2 was low in S1-S2, but increased in S3-R. Therefore, the induction of cell death molecules during early onset might be associated with lymphocyte and NK cells death in S2. We found that ARG1 (arginase) had an increase in S2 (Fig. 5D). Arginase is constitutively expressed by neutrophils and is an enzyme that metabolizes arginine into ornithine and urea. The depletion of arginine may impair T cell response. Furthermore, we also found that LAG3, TIM-3 (HAVCR2), and transcription factors (EOMES, NFATC1, NFATC2, PBX3, and FOXO3) that associates with T cell exhaustion, increased in S1-S2 (Fig. 4C, Fig. S4A).

**Fig. 5.**
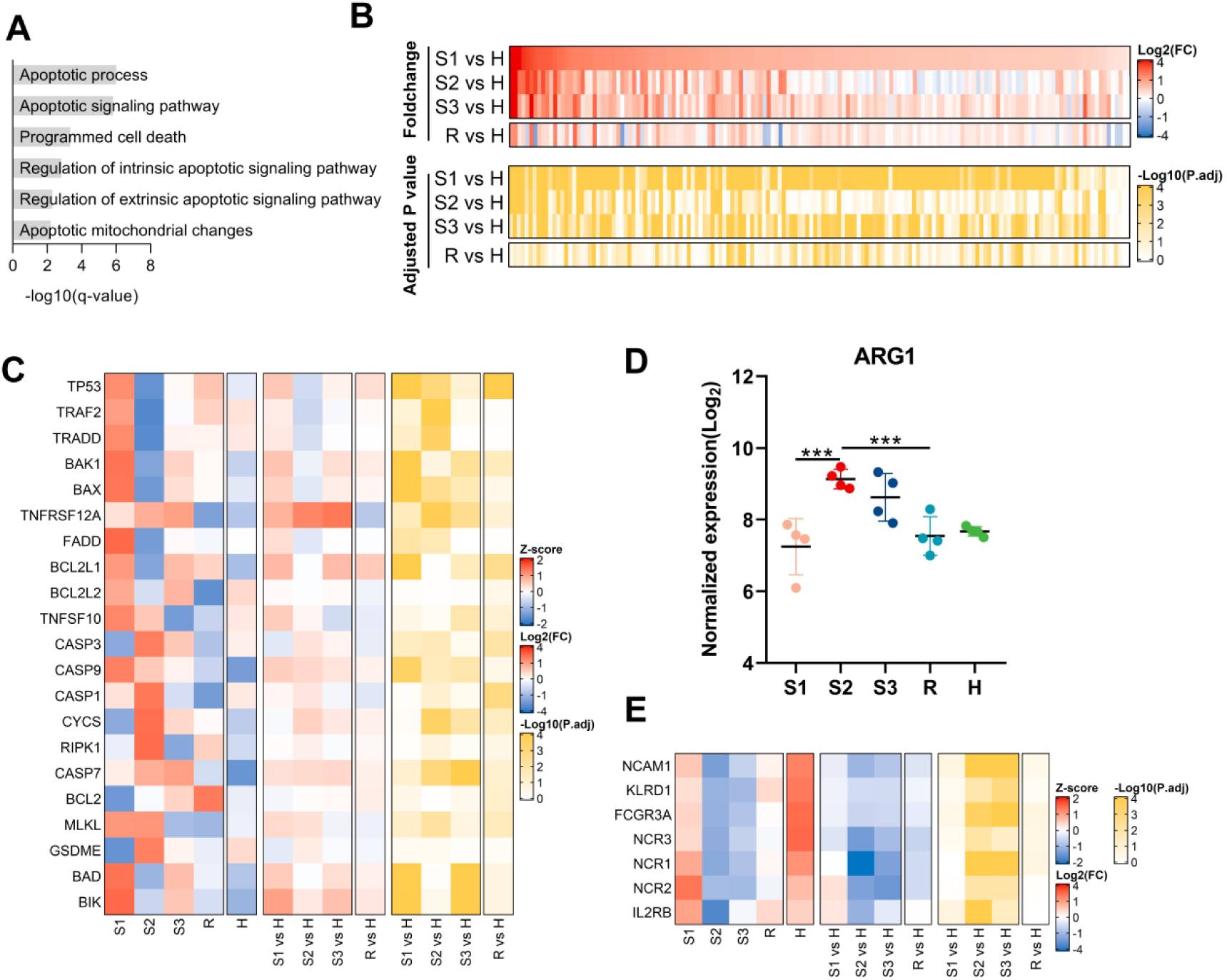
Reduction of lymphocytes and NK cells are caused by dysregulated activation of cell death, exhaustion, and migration. (**A**) Go analysis of transcripts included in cluster “M” or “n” showing enrichment for cell death pathway-related biological processes. Horizontal axis denotes statistical significance as measured by minus logarithm of q-values. Vertical axis ranked the Go terms by q-values (gray bars). (**B**) Heatmaps showing foldchange (top panel) and corresponding adjusted p values (bottom panel) for cell death pathway related transcripts. (**C**) Heatmaps showing relative expression level (left panel), foldchange (middle panel) and adjusted p values (right panel) for a subset of cell death pathway associated genes. (**D**) Normalized log_2_ expression of ARG1 for each sample from individual COVID-19 patients or healthy controls. (E) Heatmaps showing relative expression level (left panel), foldchange (middle panel) and adjusted p values (right panel) for a subset of NK cell associated genes.

Interestingly, the S1PR1, S1PR2 and S1PR4 and S1PR5 which are associated with lymphocyte and NK cells egression from peripheral lymphoid organs decreased to the lowest level in S2, while the S1PR3 that mostly expresses in monocytes increased in S2 (Fig. S5B-C). NK cell associated genes, including NCAM1 (CD56), FCGR3A (CD16), KLRD1 (CD94), NCR1, NCR2, NCR3, and IL2RB, decreased in S1-S2 but resumed in S3-R (Fig. 5E, Fig. S5D), indicating a reduction of NK cells during early onset. These results suggested that multiple factors including cell death and migratory molecules, collectively contribute to the reduction of peripheral T lymphocytes and NK cells.

### SARS-CoV-2 infection induced coagulation disorders which may be exacerbated by hypoxia

We observed that blood coagulation related genes were significantly enriched in cluster “n”, including blood coagulation, platelet activation, plasminogen activation, blood vessel development, fibrin clot formation (Fig. 6A-B). Coagulation factors (F3, F5, F12, and F13A1) and platelet-activating genes (GP1BA, GP1BB, and GP9) were elevated in S1 and maintained at a high level until S3. Several transcripts related to the process of fibrin dissolution, including PLAU (urokinase plasminogen activator, uPA) and PLAUR (uPA receptor), increased during S1-S3 (Fig. 6C-D, Fig. S6). PLAU and PLAUR acts to convert plasminogen to plasmin, which promotes fibrinolysis, leading to generation of D-dimers (fibrin degradation products). D-dimer is a known hemostasis marker that reflects ongoing fibrin formation and degradation. In clinical tests, the 4 patients in the study had average D-dimer level at 725 ± 371 μg/L in S1, which was much higher than that in healthy people (45 ± 23 μg/L) (Fig. 6E). Paradoxically, SERPINE1 which encodes plasminogen activator inhibitor type 1(PAI1), also elevated during S1-S3. PAI1 inhibit tissue-type plasminogen activator (tPA) and uPA. Moreover, additional serpin family members (SERPINA1, SERPINB2 and SERPING1) also showed elevated expression during S1-S3 (Fig. 6C, Fig. S6). We speculate that PAI1 may inhibit uPA and tPA mediated fibrinolysis and stimulate thrombus formation in COVID-19 patients. These data suggest that coagulation disorder occurs early and lasts most period of COVID-19 disease progression.

**Fig. 6.**
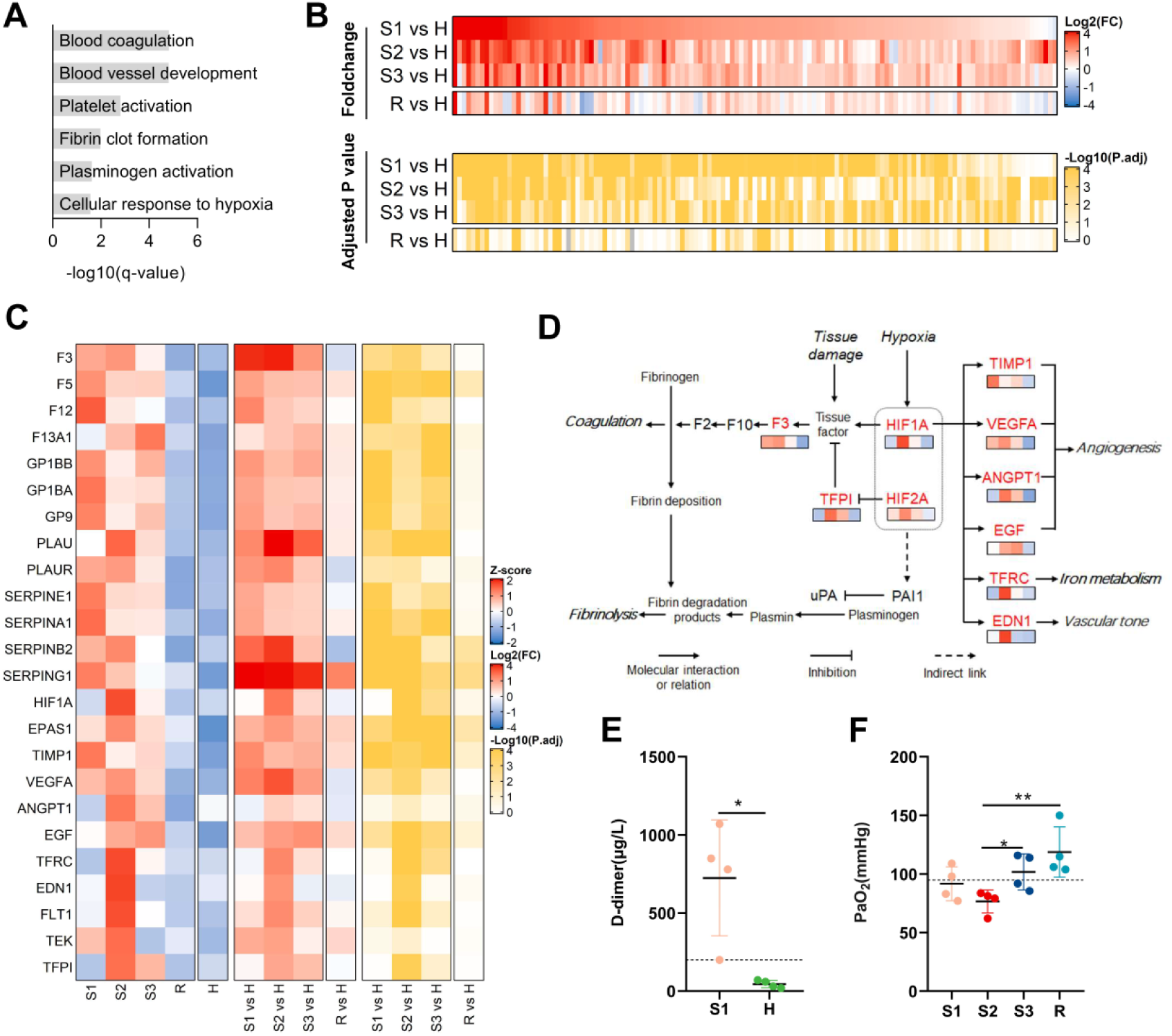
SARS-COV-2 infection induced coagulation disorders and hypoxia. (**A**) Go analysis of transcripts included in cluster “n” and “A” showing enrichment for coagulation and hypoxia-related biological processes. Horizontal axis denotes statistical significance as measured by minus logarithm of q-values. Vertical axis ranked the Go terms by q-values (gray bars). (**B**) Heatmaps showing foldchange (top panel) and corresponding adjusted p values (bottom panel) for coagulation and hypoxia-related transcripts. (**C**) Heatmaps showing relative expression level (left panel), foldchange (middle panel) and adjusted p values (right panel) for a subset of coagulation and hypoxia associated genes. (**D**) Schematic representation coagulation, fibrinolysis and hypoxia pathways. (**E**) Plasma levels of D-dimer among healthy controls and COVID-19 patients in S1. *, p<0.05 by paired t test. (**F**) Arterial oxygen partial pressures in COVID-19 patients in four stages. **, p<0.01; *, p<0.05 by paired t test.

We found a subset of genes involved in hypoxia response was enriched in cluster “A” (Fig. 6A-B). HIF1A and EPAS1 (HIF2A) are the key genes responding to low oxygen, were significantly elevated in S2, although the hypoxia was likely occurred in S1 (Fig. 6C, Fig. S6). Its downstream genes, including VEGFA (vascular endothelial growth factor A), EGF (epidermal growth factor), ANGPT1 (angiopoietin1), TIMP1 (TIMP metallopeptidase inhibitor1), EDN1 (Endothelin1), and TFRC (transferrin receptor) increased in S2 and/or S3 (Fig. 6C, Fig. S6). The upregulation of these genes contributes to angiogenesis, accelerated iron metabolism, and vascular contraction, which can help to improve oxygen delivery. Indeed, clinical observation of the patients in this study showed their arterial oxygen partial pressures started to decline in S1 and were the lowest in S2 (Fig. 6F).

Notably, hypoxia can induce SERPINE1 expression and thus further enhances thrombosis (*18*). HIF2A represses the expression of TFPI (tissue factor pathway inhibitor) to release the inhibition on TF (tissue factor). HIF1A facilitates TF expression, which triggers the extrinsic pathway of the coagulation cascade (*19, 20*). Hypoxia thus may further disrupt the homeostasis of fibrin degradation. These factors in combination may exacerbate disorders in blood coagulation, resulting in disseminated intravascular microthrombosis in COVID-19 patients.

### Potential biomarkers in PBMCs transcripts for predicting COVID-19 progression and prognosis

We applied multi-category classification (MCC) based on logistic model to identify potential biomarkers in transcriptome level. All PBMCs transcriptomes were used as a “discovery set”. Multi-class ROC (Receiver Operating Characteristic curve) followed by AUC (Area under the Curve of ROC) calculation were performed to assess diagnostic accuracy (*21*). We identified 25 genes with the highest AUC from cluster “n”, “A”, and “M”: IL6, IL10, CXCL8, CXCL2, ALOX15B, LRG1, IL1R2, IL1RN, MMP9, GRP84, OASL, MX1, CD3E, CD3G, CD8B, TP53, BAX, BAK1, IGLL5, S1PR1, F3, PLAU, SERPINB2, SERPINE1, and HIF1A. These genes exhibited highly conserved kinetics among different COVID-19 patients (Fig. S7A). The PCA analysis using the combination of these 25 genes was sufficient to define different disease stages without using the whole transcriptomes (Fig. 1B, Fig. S7B).

Finally, we attempted to further identify biomarkers that may predict the risk of severe disease during the early onset (S1), preferably before the change occurs for traditional biomarker CRP (which was elevated during S2 in our patients). We performed another RNA-seq analysis for PBMCs collected from 3 severe cases during their early stages (day 4, 6, and 8 after symptom onset, respectively) before these patients were transferred to ICU. 12 genes showed good predictive power of disease severity prior to or parallel to clinically severe manifestation were revealed, including IL6, IL10, CXCL2, ALOX15B, IL1R2, MMP9, GPR84, F3, SERPINE1, CD8B, CD3G, and S1PR1 (Fig. 7A). AUC analysis based on the expression of these 12 genes showed an excellent prediction of the severity (Fig. 7B). This result demonstrated that the combinatorial analysis of 12 genes could help to predict if a patient has a high risk of developing severe disease even when the disease is in its early course.

**Fig. 7.**
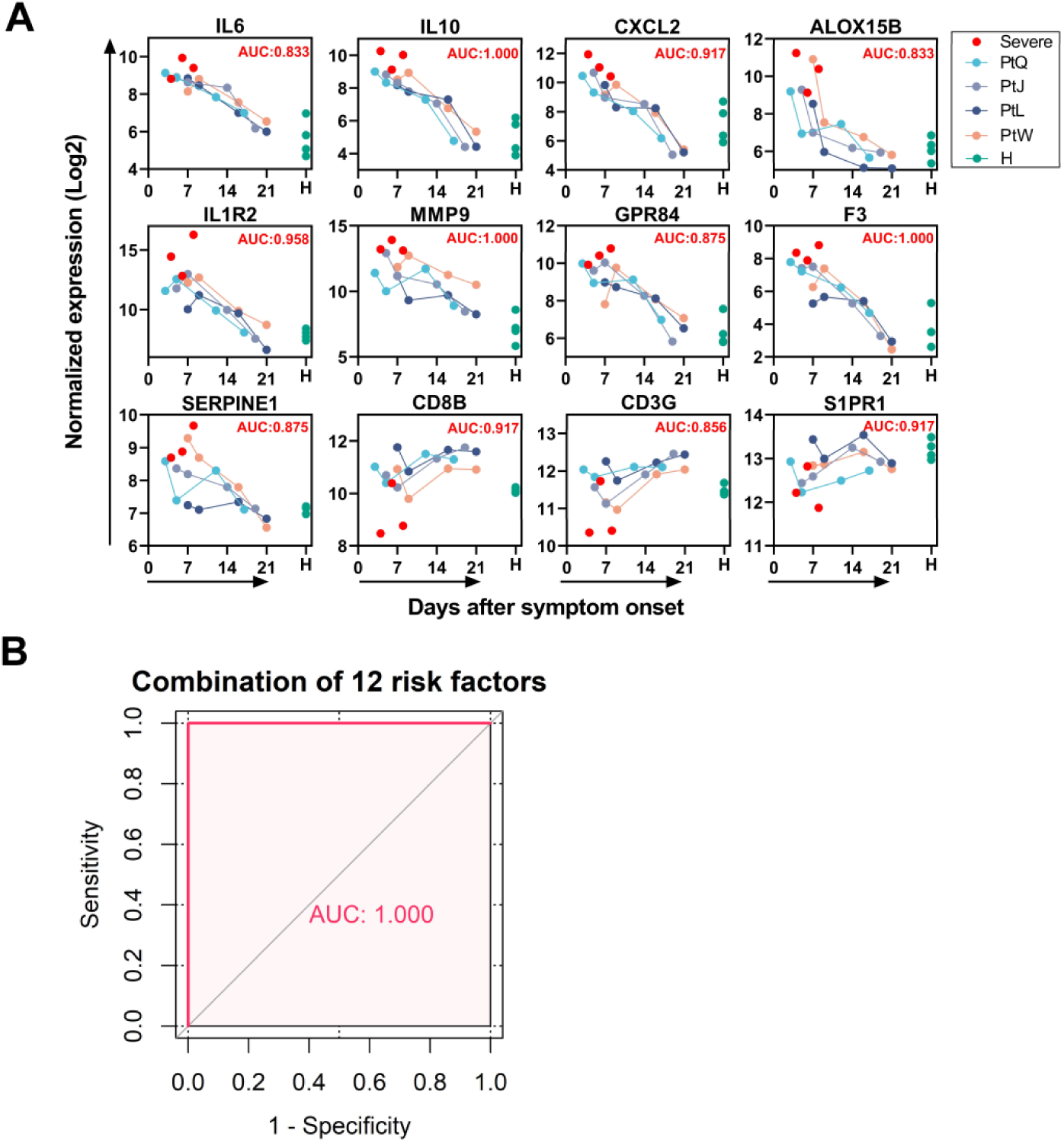
Identification of potential biomarkers. (**A**) Normalized log_2_ expression and AUC values of 12 potential biomarkers for disease severity prediction. Three samples (read dots) collected in early stage (4-8 days) from severe patients were used to explore potential risk factors. X axis denotes the days from onset on which the corresponding sample was collected. Y axis denotes log2 normalized gene expression. AUC values are listed on the top left corner of each panel. (**B**) Receiver-operator characteristics (ROC) curve of a combination of 12 risk factors for disease severity prediction.

## DISCUSSION

The worldwide spreading COVID-19 is an acute respiratory disease caused by SARS-CoV-2 infection. To date, there are no designated drugs and effective clinical treatment for COVID-19 due to the lack of understanding of host immune response to SARS-CoV-2 infection. To better control disease progression and minimize fatality, it is important to identify patients who may progress into severe or critical condition; therefore, appropriate counter-measures may be prepared in advance. Here, we longitudinally analyzed transcriptomes of PBMCs collected from COVID-19 patients. We found SARS-CoV-2 infection caused dysregulated expression of inflammatory cytokines and lipid mediators and aberrant increase of pathological low-density neutrophil, leading to excessive inflammation and tissue damage. Multiple pathways, including cell death, exhaustion and migration, contributed to lymphocytes reduction and perturbation in adaptive immune responses. SARS-CoV-2 infection induced disordered coagulation and hypoxia, which exacerbate disseminated intravascular microthrombosis in COVID-19 patients. Based on our analysis, we identified and proposed 12 genes as markers that may be used to predict disease progression, severity, and prognosis.

COVID-19, SARS, and MERS, the three respiratory diseases caused by coronavirus infection, all exhibit excessive inflammatory response (*3, 22*). The inflammatory response in patients with COVID-19 may be related to disease progression and severe outcomes (*3, 23*). We and others have demonstrated that plasma levels of IL-6 and IP-10 (CXCL10) in COVID-19 patients are significantly increased (*3, 12*). In our study, the mRNA expression of cytokines IL1B and IL6 in PBMCs is also increased. These two cytokines are the key regulator of the immune system and promote the production of a large number of inflammatory factors through the JAK-STAT signaling pathway. It has been suggested that STAT-JAK pathway inhibitors and cytokine-targeted monoclonal antibodies (such as α-IL-6, etc.) have the potential to treat COVID-19 (*12, 23, 24*). In this study, lymphocyte counts were reduced to the lowest level in stage S2 in COVID-19 patients. At the same time, the expression of IL2, IL7, IL15 and IL21 is increased, which may promote the proliferation of lymphocytes and facilitate the recovery of their number and function (*25-27*).

We observed the increase of enzymes involved in synthesis of lipid mediators. Pro-inflammatory lipid mediators such as PGE2 may exacerbate inflammatory response in COVID-19 patients. PGE2 as a pro-inflammatory mediator can disrupt the balance of Th1 and Th2 (*28*). Previous reports demonstrated that PGE2 can inhibit the production of IFNα and β via blockage of PI3K/AKT pathway (*29-31*). This may partially explain why the expression of type I IFNs was undetectable in PBMCs of COVID-19 patients. Interestingly, we also observed an increase of ALOX15B. It has been reported that PGE2 can induce ALOX15 expression through the receptor EP4 signaling pathway and promotes the production of the anti-inflammatory mediator LXA4 (*32*). Indeed, the gene expression finding was verified by observing an increase of plasma LXA4 in patients. It has been proposed that the use of non-steroidal anti-inflammatory drugs such as COX-2 inhibitors (e.g. Celecoxib) to inhibit the production of prostanoids may reduce inflammation (*33*). Therefore, the appropriate time and dosage of using NSAIDs in COVID-19 patients should be carefully evaluated.

In this study, we observed the expression of NET-related genes was increased in COVID-19 patients. As a key pathogenic factor of neutrophils, NETs play a key role in the initiation and persistence of autoimmune responses and tissue damage (*34*). NETs are closely related to the severity of influenza, Ebola virus disease, and dengue hemorrhagic fever. In severe patients with influenza virus infection, excessively activated neutrophils form excessive NETs (*35*). In Ebola virus disease fatalities, excessive NET-related proteins (MPO, CTSG) expressed by neutrophils are closely related to systemic tissue damage caused by Ebola virus infection (*36*). In the acute phase of dengue virus infection, patients with dengue hemorrhagic fever (a more severe disease state) have higher levels of NET-related proteins (MPO-DNA complexes) (*37*). Besides, the number of neutrophils in SARS and COVID-19 severe patients is significantly increased (*1, 38-40*). Additionally, CXCL8 is secreted primarily by neutrophils. CXCL8 serves as a chemotactic factor by guiding the neutrophils to the site of infection to cause lung damage.

The IFNs play an essential role in the antiviral response, but viruses can also escape from host immunity by inhibiting type I interferons. Although the upstream signaling pathway of IFNA/B was activated in PBMCs of all four COVID-19 patients, few IFNA/B transcripts could be detected, suggesting that the virus might suppress the transcription of IFNs. Besides the possible impact of pro-inflammatory lipid mediators, the detailed mechanism needs to be further investigated. It has been reported that the expressions of IFNs were induced by influenza virus but not SARS-CoV (*41*). The West Nile virus (WNV) NS1 protein and Kunjin virus NS2A protein inhibited the activation of IRF7 and the transcription of IFNB, respectively (*42*). However, ISGs were significantly up-regulated. We could not exclude the possibility that type I interferons were induced very early before the symptom onset and then suppressed by the time these samples were collected. It is worthwhile to notice that the increased expression of IFN-γ in these patients was consistent with the severity of the disease. The IFN-γ also significantly increased in severely patients with SARS-COV, MERS-COV as well as H7N9 infection (*11, 43, 44*). We speculated that IFN-γ might induce large amounts of cytokines secretion to aggravate the illness. Therefore, the drugs that lower the expression of IFNγ may help to alleviate the disease severity.

SARS-CoV-2, like SARS-CoV, MERS-CoV, and H5N1 virus, also induce the reduction of peripheral blood lymphocytes (*45*). Although hperactivated and exhausted T cells undergo apoptosis and contribute to cell number reduction (*46-48*), we found that there might be other factors involved in the reduction of peripheral blood lymphocytes in COVID-19 patients. Firstly, besides the induction of intrinsic apoptotic pathways, extrinsic apoptosis, necrosis and pyroptosis associated molecules also elevated in S2. Secondly, coincided with our findings, Ebola virus infection also elevates neutrophil number and arginase expression, resulting in peripheral T lymphocytes to undergo neutrophil-mediated and arginase-dependent cell number reduction (*36*). Depletion of cellular arginine can also induce apoptosis of T cells (*49*), suggesting arginase increase may also contribute to the reduction of lymphocytes in COVID-19 patients. Currently, arginase inhibitor INCB001158 has been used in Phase I / II clinical trials for the treatment of patients with advanced metastatic solid tumors (NCT03314935), so this inhibitor may be considered for the treatment of COVID-19 patients. We propose an even simpler solution by supplementing arginine to COVID-19 patients that may help to recover from lymphopenia. Finally, we found that the four S1PR receptors decreased to the lowest level in S2, which may reduce the egression of T and NK cells from peripheral lymphoid organs to the blood (*50, 51*).

In contrast, S1PR3, which is mainly expressed by monocytes, had increased expression in S2. The inhibitors of S1PR1 and S1PR5, Fingolimod (FTY720, trade name Gilenya) and Siponimod (trade name Mayzent) are being used to treat multiple sclerosis (MS). These two drugs should be used with caution if the patient is infected with SARS-COV-2.

Some COVID-19 patients developed disordered coagulation, including disseminated intravascular coagulation (DIC) (*7, 52, 53*). We found that COVID-19 patients had increased expression of many coagulation associated genes started from early-onset and lasted two-third of the disease course. The patients in this study also had increased D-dimer in the serum, indicating fibrin deposition. As a consequence, PLAU and PLAUR were elevated for fibrin degradation. However, it was reported that 71.4% of the non-survivors matched the grade of DIC in the later stages of COVID-19 (*52*). Two recent reports showed that COVID-19 patients had pulmonary thrombosis and bleeding lesions (*54, 55*). We found that the fibrin degradation pathway is disrupted in severe ways. The expression of SERPINE1 (PAI1) and its family members are significantly increased during early onset. PAI1 stimulates thrombus propagation by inhibiting uPA (PLAU) mediated fibrinolysis. Moreover, hypoxia upregulates expression of HIF1A and HIF2A which further suppress uPA (PLAU) activity, leading to secondary hyperfibrinolysis condition. Previous study suggested that dysregulation of the urokinase pathway during SARS-coronavirus infection contributes to more severe lung pathology (*56*). Our study suggested that SERPINE1 and PLAU may be key targets involved in the early pathological process of DIC. There may be other causes of coagulation disorders in COVID-19 patients. For example, LDN releases excess neutrophils extracellular traps (NETs) that cause platelets to accumulate at the site of inflammation and promote thrombosis (*57*). It is particularly important to monitor coagulation indicators in the COVID-19 patients closely and actively promote the relevant methods of coagulation diagnosis in COVID-19 patients.

We identified 25 gene expression markers that can be used to identify the disease progression. Importantly, combinatorial analysis of expression of 12 genes may be able to predict if a patient is at high risk of progressing to severe disease, even when the patient shows no clinical signs of severity. Due to limited early-stage PBMC samples from patients who later ended up in ICU, future studies should be carried out by investigators who can obtain more samples to validate and improve this gene set for early identification, therefore improve outcomes in high-risk patients

## MATERIALS AND METHODS

### Study design

This study was designed to understand the host pathophysiological responses after SARS-CoV-2 infection. Peripheral blood mononuclear cells were collected from 4 COVID-19 patients at 4 different time points from symptom onset to recovery. For comparison, 4 blood samples from a healthy donor before and after vaccination with a QIV inactivated seasonal influenza virus vaccine were used as healthy controls. Global expression of RNA in these samples was measured by RNA-sequencing. RT-PCR for a set of common genes was performed to exam transcriptome data validity.

### Human Subjects and Ethics

Peripheral blood mononuclear cells (PBMCs) from 4 female COVID-19 patients were obtained from Guangzhou Eighth People’s Hospital of Guangzhou Medical University. All patients signed informed consent for this study. This study was approved by Review Committee of Guangzhou Eighth People’s Hospital of Guangzhou Medical University. All healthy control subjects had written informed consent prior to the collection of peripheral blood. PBMCs were performed on existing samples collected during standard diagnostic tests, posing no extra burden to patients.

### PBMC Isolation and Total RNA Extraction

Whole blood samples were centrifuged at 3000 RPM for 15min to collect plasma. An equal volume of 1640 medium was mixed with blood cells, and peripheral blood mononuclear cells (PBMCs) were isolated with Opti-Prep lymphocyte separation solution (Axis Shield Poc As, Oslo, Norway) by following the manufacturer’s instructions. Total RNA of PBMCs was extracted using TRIzol™ (Life Technologies) according to the manufacturer’s instruction Invitrogen.

### Quantitative Real-Time PCR

Reverse transcribe RNA into cDNA using the iScript cDNA synthesis kit (#1708891, BIO-RAD). The cDNA then served as templates for quantitative RT-PCR and were amplified using a Bio-Rad CFX96 Real-time PCR Detection System (Bio-Rad) with ChamQ SYBR qPCR Master Mix (Q311-03, Vazyme). Cycle threshold [C(t)] values and melting curves were analyzed with CFX Manager 3.1 (Bio-Rad). Primers for CD4: forward, 5’-TGCCTCAGTATGCTGGCTCT, reverse, 5’-GAGACCTTTGCCTCCTT GTTC. Primers for CD8A: forward, 5’-TCCTCCTATACCTCTCCCAAAAC, reverse, 5’-GGAAGACCGGCACGAAGTG.

### Multiplex cytokines determination by Luminex assay

The plasma of COVID-19 patients was inactivated at 56 °C for 1.5 h. Multicytokines were measured using cytokine and chemokine magnetic bead panel kit (HCYTMAG-60K-PX38, Millipore), following the manufacturer’s instructions.

### Blood Biochemistry Detection

Venous blood was collected from COVID-19 patients and serum was isolated according to conventional methods. The levels of C-reactive protein (CRP) and blood urea nitrogen (BUN) in the serum were determined by automatic biochemical analyzer (CL8000, SHIMADZU, Japan).

### RNA-seq Library Construction and Sequencing

Extracted RNA was DNase treated with 1 U of Baseline Zero DNase (Epicentre) at 37 °C for 30 min, cleaned with 1.8X volume of AMPureXP beads (Beckman-Coulter), and eluted in nuclease-free water. RNA quality was assessed using an Agilent Bioanalyzer (all samples exhibited RNA integrity numbers > 9) and quantified using the Qubit RNA Broad Range Assay kit (Thermo Fisher). 200ng of each DNase-treated sample was used for library preparation. Briefly, ribosomal RNAs were depleted using the (QIA seq FastSlect-rRNA HRM KIT, QIAGEN) according to the manufacturer’s instructions, Ribosomal RNA depletion was confirmed by using Agilent Bioanalyzer analysis and noting the absence of ribosomal peaks. Next, Prime, and Fragment Mix from the (NEB Next RNA library prep Kit) were added to each sample, followed by fragmentation at 94 °C for 8 min to yield a median fragment size distribution of 155 nt and a final library of 309 nt. Libraries were prepared according to the manufacturer’s instructions by using the NEB Next RNA Library Prep Kit, incorporating different barcoded adaptors for each sample and amplifying libraries for 15 cycles. Following final library quality control on the Agilent Bioanalyzer to confirm the expected size distributions, libraries were pooled and sequenced on the Illumina HiSeq3000 platform in a 150-bp paired-end read run format.

### Pre-Processing of the Raw RNA-seq Data

Raw RNA-seq reads were filtered according to their base qualities, read sequences were trimmed at 3’end after reaching a 2-base sliding window with PHRED quality score lower than 20. Following filtering, Illumina adapter sequences at 3’end were removed using Trimmomatic v0.36 (*58*). After low-quality filtering and adapter trimming, reads more than 50 nt in length were retained for further analysis. Next, the trimmed reads were mapped to the human (hg38) and SARS-COV-2 viral (Wuhan-Hu-1) reference genomes (*3*) using HISAT v2.1 (*59*) with corresponding gene annotations (Gencode GRCh37/V32 for the human genome) with default settings RF, respectively. Total counts per mapped gene were determined using featureCounts function in SubReads package v1.5.3 (*60*) with default parameter.

### RNA-seq Data Analysis

Raw counts matrix obtained from featureCounts was used as input for differentially expression gene analysis with the bioconductor package edgeR v3.28 (*61*) or DESeq2 v1.26 (*62*) in R v3.6. Gene counts more than 5 reads in a single sample or more than 100 total reads across all samples were retained further analysis. Package edgeR was used to perform paired comparison between single samples (Fig. 2A and Supplementary Fig. 2A). Normalization factors were computed on the filtered data matrix using the weighted trimmed mean of M-values (TMM) method, followed by voom mean-variance transformation in preparation for Limma linear modeling. Differential expression gene analysis was enlisted exactTest function and the biological coefficient of variation (BCV) parameter was set to 0.4. Genes with log2 fold change >1 or <−1 and false discovery rate (FDR) p-value < 0.05 were considered significant. The same counts matrix was used for pairwise group comparison (Fig. 2C and Supplementary Fig. 2C), and normalized using the DESeq2 method to remove the library-specific artefacts. Genes with log2 fold change >1 or <−1 and adjusted p-value <0.05 corrected for multiple testing using the Benjamini–Hochberg (BH) method were considered significant.

### Global Transcriptome Analysis and Digital Cytometry

To determine the similarity between the PBMCs sample, the normalized counts matrix was used to calculated pairwise person coefficients using build-in function cor in R software. Hierarchical clustering across all samples was based on pairwise Pearson correlation coefficients matrix. And the build-in function prcomp in R software was enlisted for principal component analysis. We inferred the immune cell quantities in each blood sample using the CIBERSORT server (https://cibersortx.stanford.edu/).

### Timeseries-Based Gene Expression Pattern Analysis and Gene Ontology (GO) Enrichment Analysis

To explore the gene expression pattern of the DEGs, we applied the R package Mfuzz v2.46 (*63*)for time-series analysis. The healthy samples were assumed to be a stage representing pre-infection. We first run fuzzy c-means clustering algorithm for a range of c values and compare the results. The minimum distance D.min between cluster centroid was utilized as a cluster validity index. Finally, we chose c = 6 as an optimal cluster number and all the DEGs were divided into 6 clusters according to their expression pattern. subsequently, gene ontology analysis was performed to assess their biological relevance using R package clusterprofiler v 3.14.3 (*64*).

### Determination of Expression and Diversity of TCR and BCR in RNA-seq Data

We determine the expression of rearranged TCR and BCR in RNA-seq samples using MiXCR v3.0.3 (*65*). MiXCR is a universal tool for fast and accurate analysis of T-and B-cell receptor repertoire sequencing data, which also provides parameter -p rna-seq for processing RNA-seq data. Here, we aligned the filtered reads (see in Pre-processing of the raw RNA-seq data) against reference V(D)J genes that download from IMGT (http://www.imgt.org/). And the total matched counts of TCR or BCR of each sample were normalized according to their sample size factors. To analysis the diversity of TCR and BCR, we defined a clone as group of sequences with the same VH gene, JH gene and identical CDR3 amino acid sequence. The clonal diversity of TCR and BCR were analyzed using the R package Alakazam v0.3.0 (*66*). Alakazam provides an implementation of the general diversity index (*qD*) proposed by Hill (*67*), which includes a range of diversity measures as a smooth curve over a single varying parameter q. Special cases of this general index of diversity correspond to the most popular diversity measures: species richness (*q* = 0), the exponential Shannon-Weiner index (as *q→*1), the inverse of the Simpson index (*q* = 2), and the reciprocal abundance of the largest clone (as *q→∞*).

### Statistical analysis

All analyses were conducted by Prism v.8 (GraphPad Software, La Jolla, CA, USA). Comparisons between groups were analyzed by unpaired Student’s t test; multiple comparisons were performed by one-way ANOVA. A value of P<0.05 was considered statistically significant.

## Data Availability

All raw RNA-seq data used in this study have been deposited at the National Genomics Data Center (https://bigd.big.ac.cn/) under the accession number: PRJCA002599. All scripts used in this study are publicly available at https://github.com/ChenLinglab/.

## SUPPLEMENTARY MATERIALS

## Funding

This work was supported by the National Natural Science Foundation of China (82041014 and 81661148056), the Chinese Academy of Sciences Pilot Strategic Science and Technology Projects (XDB29050701).

## Author contributions

L.C. and L.Q. designed and initiated the project. Q.W and J.L. coordinated the project. C.L., X.M., J.W., F.Z. and F.L. recruited the patients. P.L., Z.C., X.Y., X.H., Y.F., T.J. and X.N. conducted the experiments. Q.Y., Y.Z., K.L., and Z.C. performed the bioinformatics analysis. L.C., L.Q., P.L., Z.C., X.Y., and X.H. analyzed the data, L.C., L.Q., Q.Y., P.L., Z.C., X.Y., and X.H. wrote the manuscript, L.C., L.Q., J.W., and L.F. revised and improved the manuscript.

## Competing interests

The authors declare no competing interests.

## Data and materials availability

**Fig. S1.**
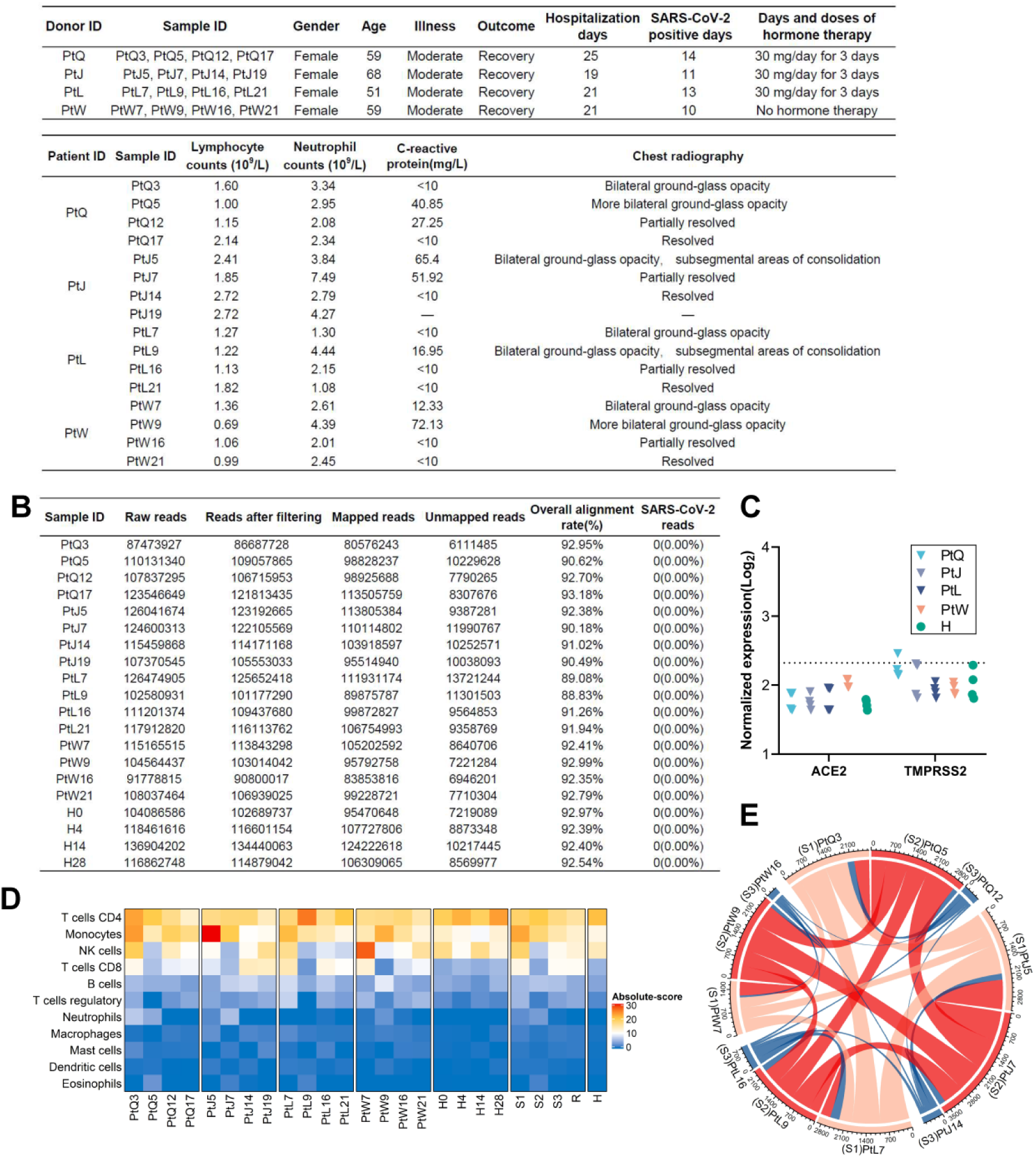
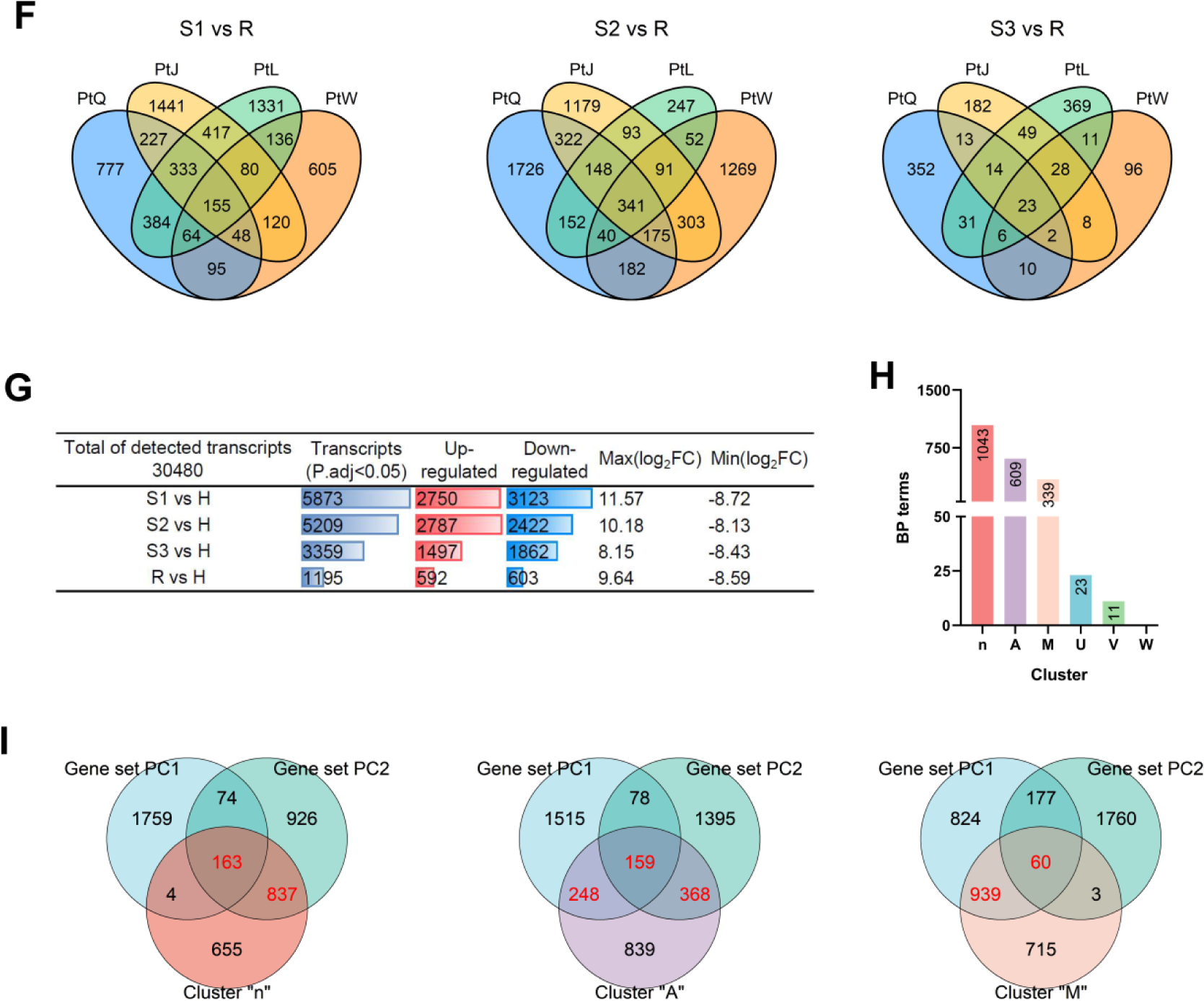
Global transcriptional analysis across COVID-19 patients and health donor samples. (**A**) Detailed information and clinical observations about each COVID-19 patient. (**B**) Summary of RNA-seq reads from PBMC samples among COVID-19 patients and healthy donor. (**C**) Normalized log_2_ expression of ACE2 and TMPRSS2 in PBMC samples. (**D**) Relative abundance of immune cell populations in PBMCs. (**E**) Circos-plot showing the overlapped DEGs between different stages in the same patient or between different patients in the same stages. In the circular plot, arcs (orange, red, and blue) are used to present S1, S2, and S3 for each patient, curved bands connecting each pair of arcs indicates the overlaps between each pair of stages, the width of the curved bands represent the number of overlapped DEGs at the two given stages. (**F**) Veen diagram showing overlapped DEGs between different patients in the same stage (left panel, S1 vs R; middle panel, S2 vs R; right panel, S3 vs R). (**G**) Samples from different patient in the same stage (S1, S2, S3, and R) were combined and compared with healthy samples (H). The number of up-regulated and down-regulated gene are listed. (**H**) Numbers of significantly enriched (q-value < 0.05) Go terms for each cluster. (**I**) Veen diagram showing overlaps and differences between genes that were contained in gene set PC1, PC2 and Cluster “n” (left panel); PC1, PC2 and Cluster “A” (middle panel); PC1, PC2 and Cluster “M” (right panel).

**Fig. S2.**
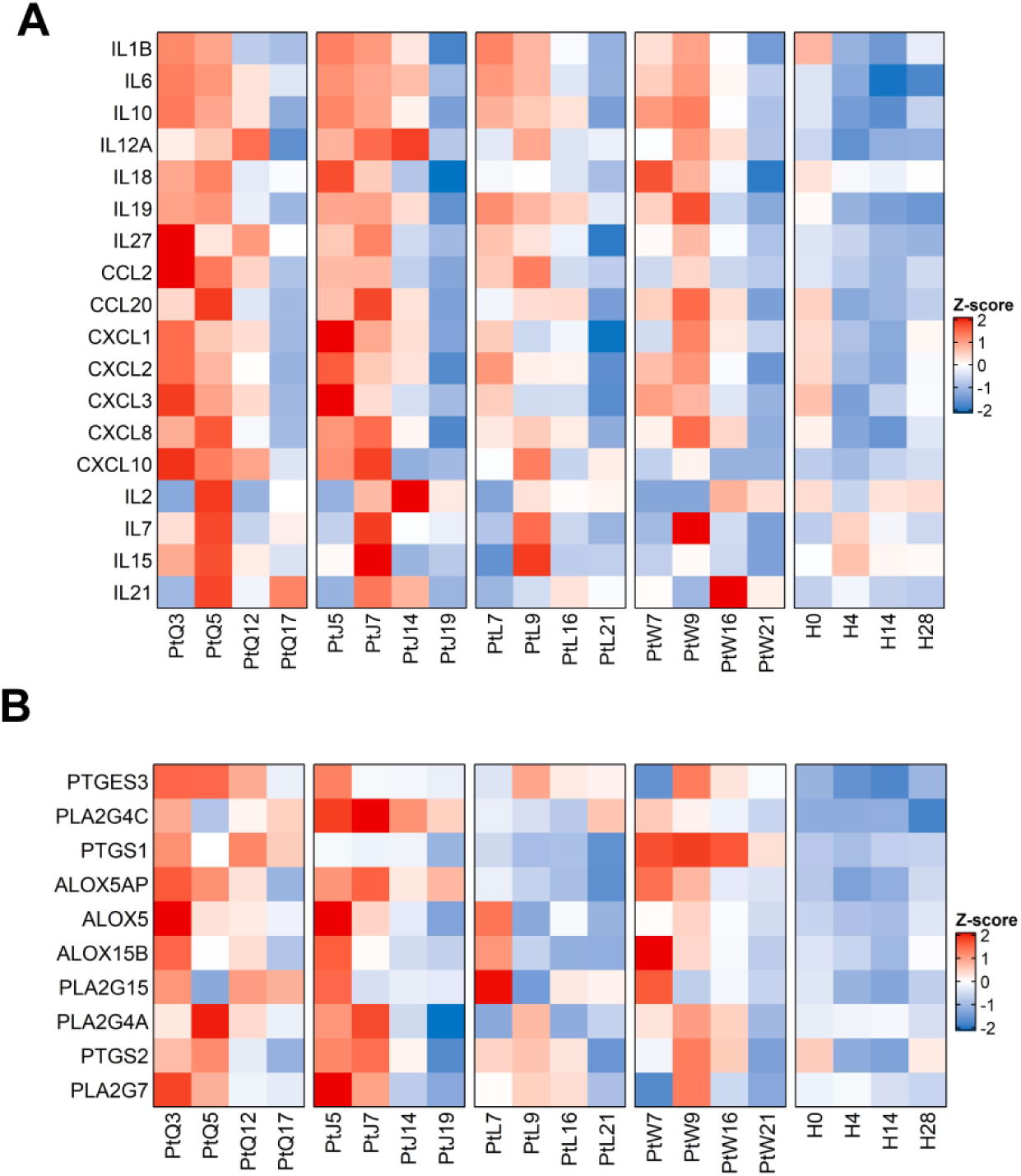
Dysregulated inflammatory cytokines and lipid mediators in COVID-19 patients. (**A**) Heatmaps showing relative expression level for a subset of inflammatory cytokines and chemokines in individual patients in four stages. (**B**) Heatmaps showing relative expression level for a subset of genes involved in prostanoids synthesis in individual patients in four stages.

**Fig. S3.**
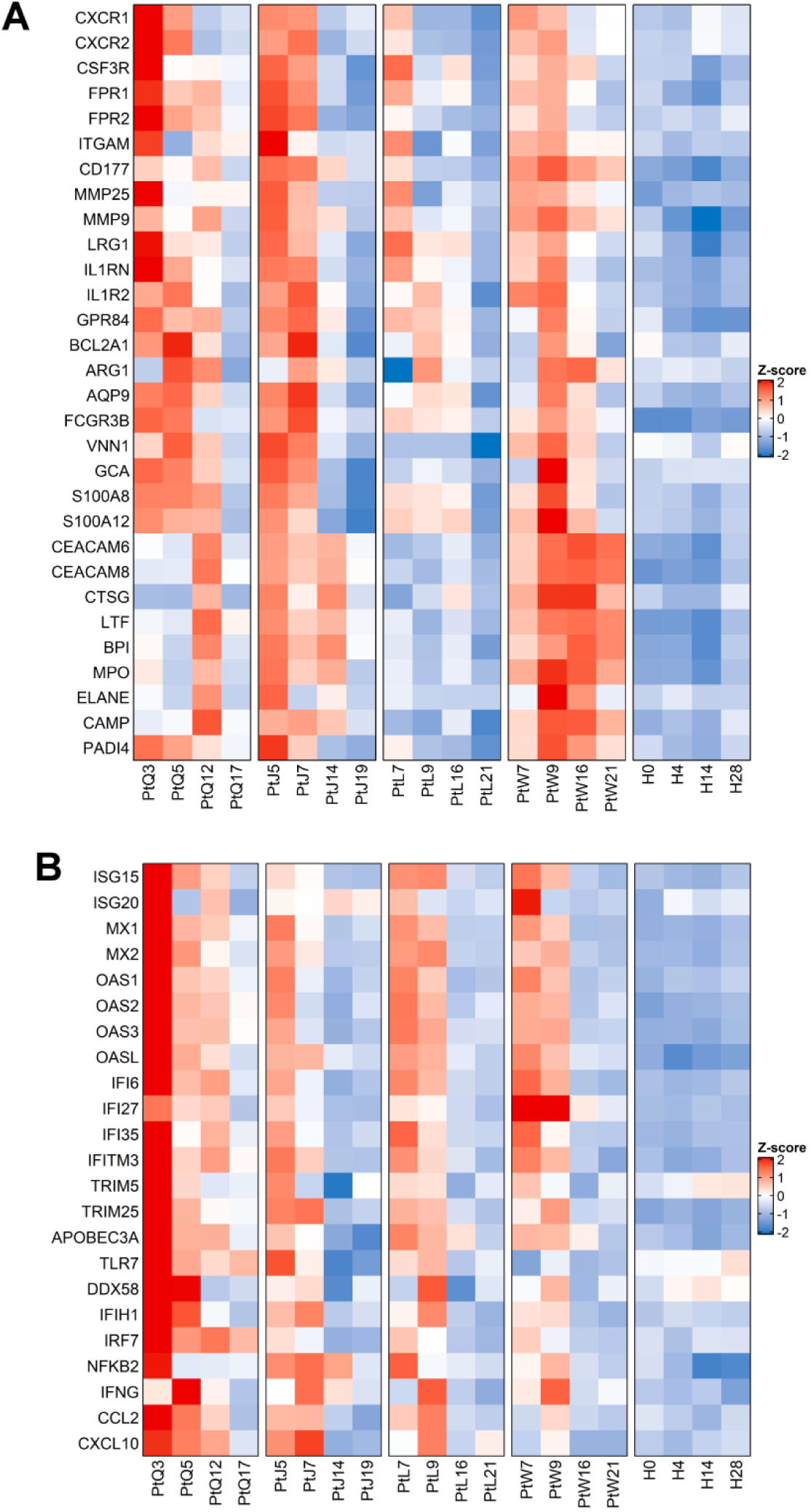
Abnormal neutrophils and dysregulated IFN responses in COVID-19 patients. (**A**) Heatmaps showing relative expression level for a subset of neutrophils in individual patients in four stages. (**B**) Heatmaps showing relative expression level for a subset of innate immune-related transcripts in individual patients in four stages.

**Fig. S4.**
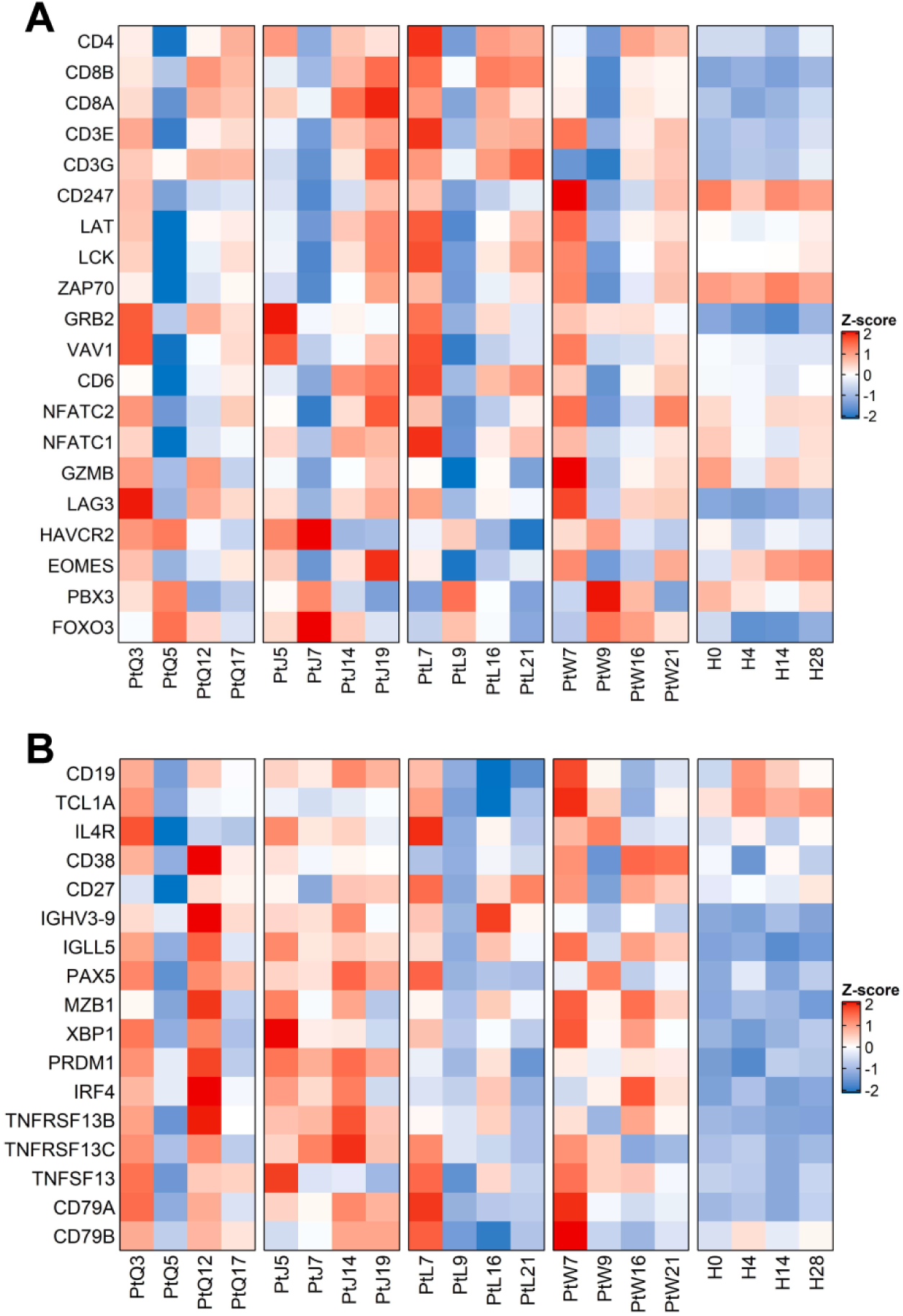
Dysregulated adaptive immune responses induced by SARS-CoV-2 infection. (**A**) Heatmaps showing relative expression level for a subset of T cell-related transcripts in individual patients in four stages. (**B**) Heatmaps showing relative expression level for a subset of B cell-related transcripts in individual patients in four stages.

**Fig. S5.**
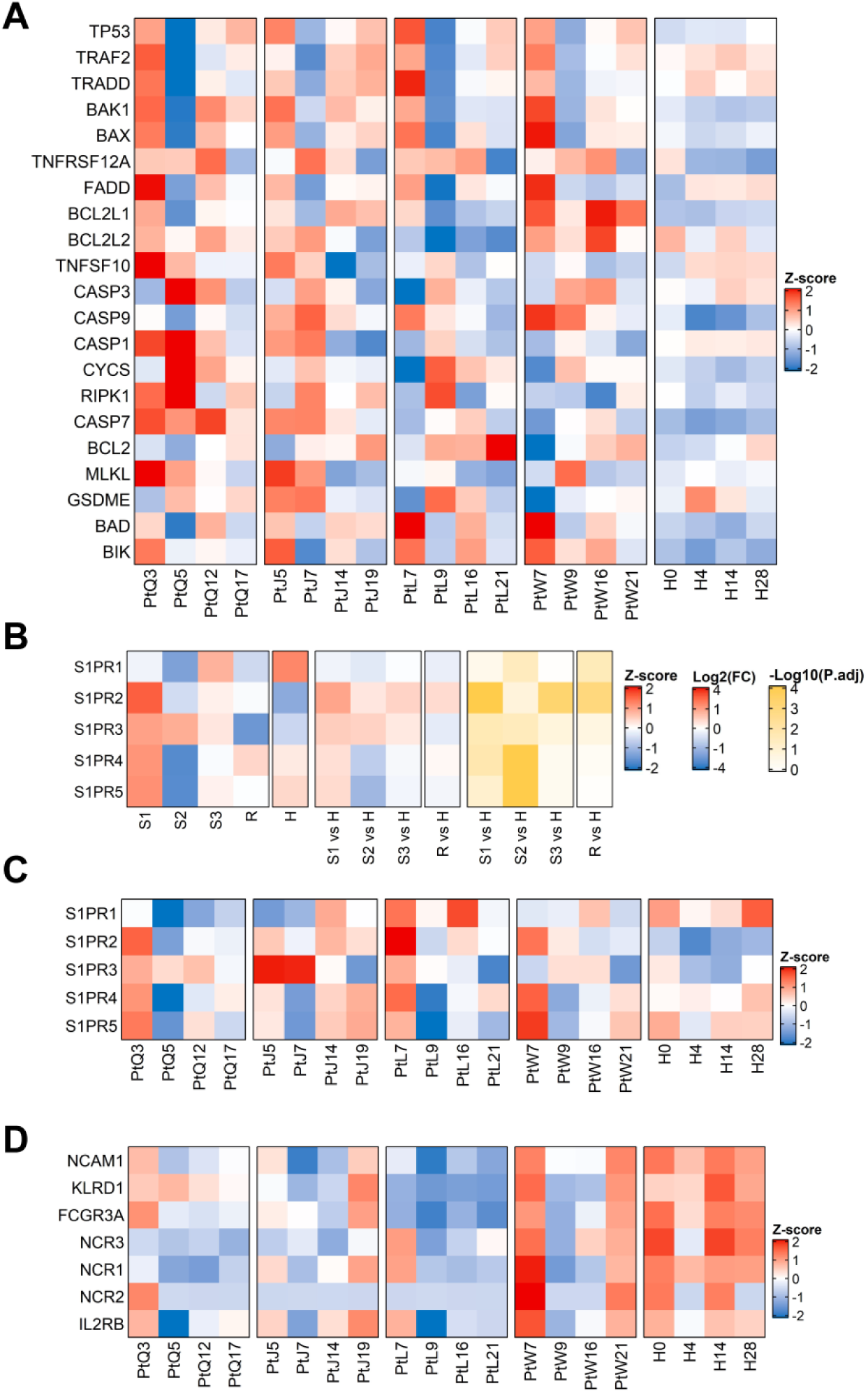
Reduction of lymphocytes and NK cells were caused by dysregulated activation of cell death, exhaustion, and migration. (**A**) Heatmaps showing relative expression level for a subset of cell death pathway-related transcripts in individual patients in four stages. (**B**) Heatmaps showing relative expression level (left), foldchange (middle) and adjusted p values (right) for S1PR1-S1PR5. (**C**) Heatmaps showing relative expression level for S1PR1-S1PR5 in individual patients in four stages. (**D**) Heatmaps showing relative expression level (left), foldchange (middle) and adjusted p values (right) for a subset of NK cell related transcripts in individual patients in four stages.

**Fig. S6.**
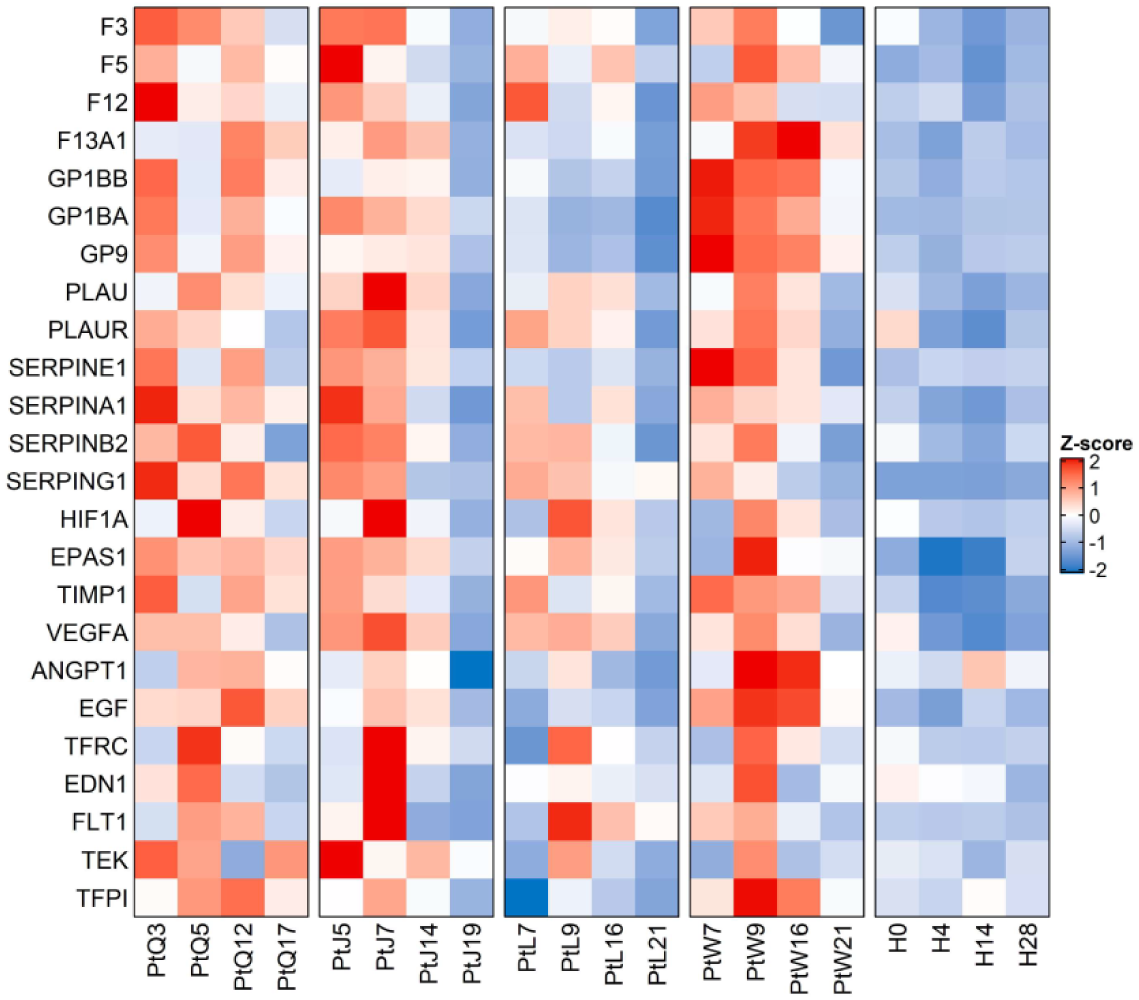
SARS-COV-2 infection induced coagulation disorders and hypoxia. Heatmaps showing relative expression level for a subset of coagulation and hypoxia-related transcripts in individual patients in four stages.

**Fig. S7.**
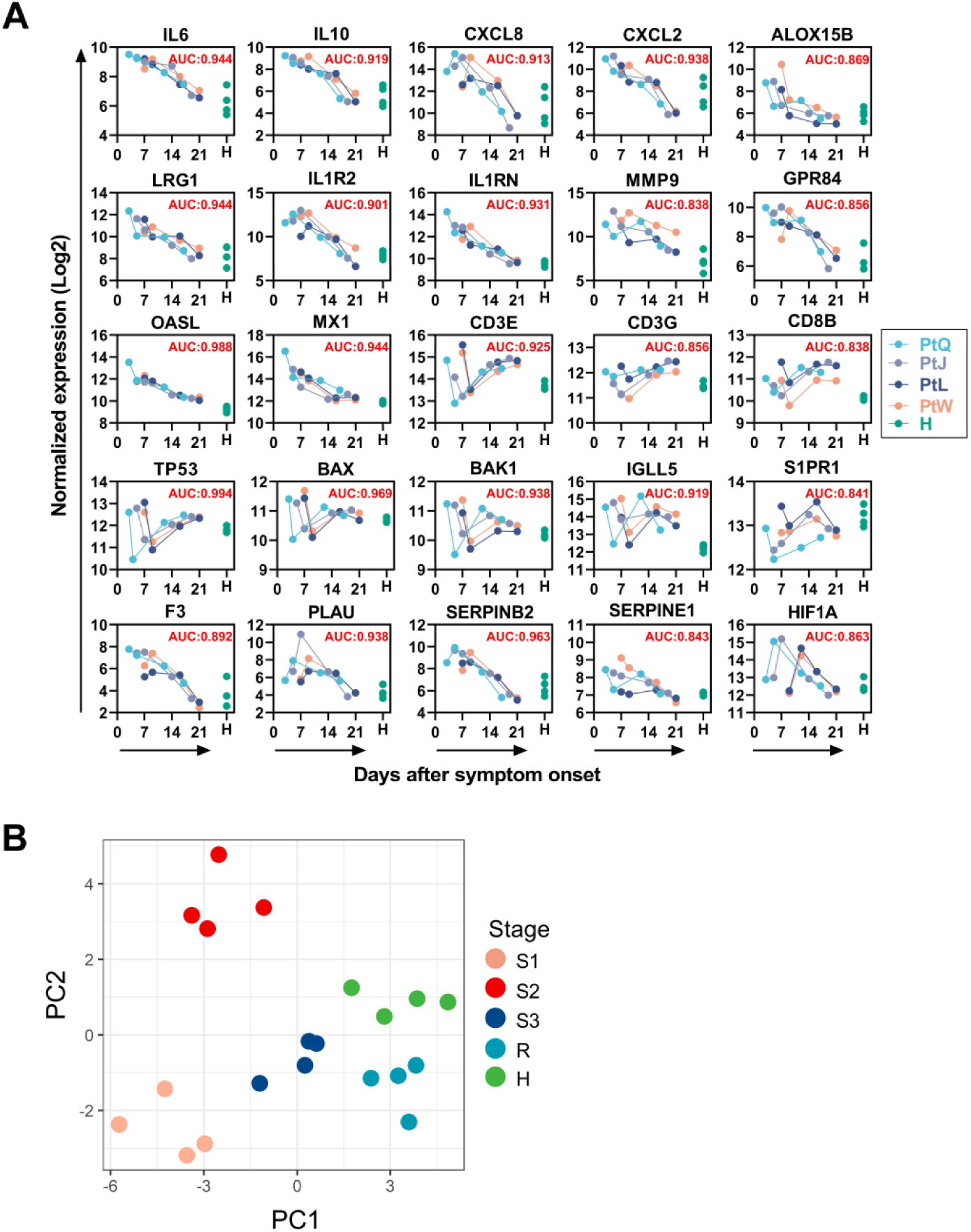
Potential biomarkers that predict outcomes of COVID-19. (**A**) Normalized log_2_ expression and AUC values of 25 candidate biomarkers for monitoring disease progression. X axis denotes the days from onset on which the corresponding sample was collected. Y axis denotes log_2_ normalized gene expression. AUC values of individual candidate biomarker are labeled on the top left corner of each panel. (**B**) Sample scores from probabilistic principal components analysis using the 25 candidate biomarkers shown in Fig. S7A.

## Notes

### Competing Interest Statement

The authors have declared no competing interest.

